# TNF blockade with certolizumab improves the efficacy of anti-PD-1 and anti-CTLA-4 combination therapy for melanoma

**DOI:** 10.64898/2026.02.11.26346073

**Authors:** Tania Margarido Pereira, Mathieu Virazels, Benjamin Jung, Thomas Filleron, Laure Badier, Etienne Leclercq, Stéphanie Brayer, Matthieu Genais, Laura Leroy, Amélie Lusque, Vincent Sibaud, Clara Maria Scarlata, Juan Pablo Cerapio, Maha Ayyoub, Muriel Mounier, Ludovic Martinet, Nathalie Andrieu-Abadie, Sergei Nedospasov, Ignacio Melero, Jean-Pierre Delord, Vera Pancaldi, Cécile Pagès, Nicolas Meyer, Céline Colacios, Anne Montfort, Bruno Ségui

**Author notes:** These authors contributed equally (Tania Margarido Pereira, Benjamin Jung). These authors contributed equally (Céline Colacios, Anne Montfort, Bruno Ségui).

## Abstract

The phase 1b TICIMEL clinical trial evaluated the safety, tolerability, and anti-tumor activity of combining the immune checkpoint inhibitors (ICI), ipilimumab and nivolumab, with tumor necrosis factor (TNF) blockers, certolizumab or infliximab, to treat advanced melanoma patients. A higher proportion of responses was observed in patients receiving ICI and certolizumab, while patients treated with ICI and infliximab demonstrated superior tolerability. Moreover, CITE-Seq analyses of circulating CD8 T cells showed that ICI plus certolizumab promoted an IFN signature, whereas ICI plus infliximab reduced the induction of genes associated with T cell activation. In preclinical models, ICI and TNF blockade with certolizumab increased IFN-γ^+^ CD8 T cells and reduced regulatory T cells in tumors. The IgG1 Fc fragment of infliximab was identified as counteracting the benefits of TNF blockade. These findings underscore the importance of selecting the optimal TNF blocker to combine with ICI to enhance therapy efficacy in melanoma patients.

ClinicalTrials.gov identifiers: NCT03293784; NCT05867004.

## Introduction

Despite the tremendous breakthrough that immune checkpoint inhibitors (ICI) such as anti-PD-1 and anti-CTLA-4 have brought to the treatment of advanced melanoma, 60% of patients do not respond or relapse within 5 years of starting treatment^1^. Moreover, ICI therapy is associated with serious immune-related adverse events (irAEs), such as colitis. This can be treated with tumor necrosis factor-alpha inhibitors (TNFi), such as infliximab^2^. The pharmacodynamic effects of TNFi on the immune response and on the clinical outcome still need to be clarified ^3,4^.

Although TNF has been identified as a soluble factor capable of inducing tumor necrosis in mice^5^, chronic production of TNF in the tumor microenvironment has been shown to contribute to cancer progression^6,7^. We and others have shown that this cancer-promoting property relies on multiple mechanisms, including immune escape. For example, TNF induces activation-induced cell death (AICD) of CD8 T cells, thereby limiting CD8 T cell infiltration into mouse melanoma tumors^8^. Thus, in mouse models of melanoma, breast and colon cancer, TNFi enhance the efficacy of ICI (anti-PD-1 in combination or not with anti-CTLA-4) and reduce the severity of irAEs such as colitis^9,10^. Therefore, combining TNF-blocking antibodies with ICI enhances therapeutic efficacy and improves tolerability *in vivo*.

Based on our preclinical studies^8,9^, we conducted a phase 1b clinical trial (TICIMEL, NCT03293784) to treat advanced melanoma patients (stage IIIc/IV) with ICI (anti-PD-1, nivolumab + anti-CTLA-4, ipilimumab) in combination with TNFi, certolizumab or infliximab. The primary objective of TICIMEL was to evaluate the safety and tolerability of the two tritherapies and the secondary objective was to assess the anti-tumor activity. While certolizumab is the monovalent Fab’ fragment of a humanized anti-TNF monoclonal antibody, linked to polyethylene glycol, infliximab is a bivalent anti-TNF chimeric IgG1 monoclonal antibody. The first part of the TICIMEL study assessed the safety on 8 and 6 patients of the certolizumab and infliximab cohorts respectively, and showed both tritherapies were safe, with a promising high response rate in the certolizumab cohort^11^. Here, we present the final results of the TICIMEL study. Tolerability was acceptable in both cohorts. Patients receiving ICI and infliximab demonstrated superior tolerability compared to those receiving ICI and certolizumab. However, most importantly, a higher proportion of patients responded to the latter. Upon attempting to explain this unexpected result, we concluded that the Fc domain of infliximab prevents TNF blockade from stimulating ICI efficacy.

We compared the effects of the two tritherapies on the protein and molecular signatures of circulating T cells from TICIMEL patients with the effects of nivolumab and ipilimumab alone, using data from a cohort of patients in the MELANFα study (NCT05867004)^12^. This uncovered that certolizumab enhances the expression of a set of genes associated with CD8 T cell activation and promotes the expression of an IFN-γ signature upon ICI. In contrast, infliximab has the opposite effect, and inhibits the ICI-dependent promotion of IFN-γ production by CD8 T cells. We also evaluated the impact of blocking TNF in combination with anti-PD-1 and anti-CTLA-4 in preclinical melanoma models using wild-type and humanized TNF and TNFR2 mice. The results demonstrate that TNF blockade improves the efficacy of the bitherapy associating anti-PD-1 and anti-CTLA-4. However, certolizumab, but not infliximab, enhanced the therapy’s efficacy by activating conventional CD4 and CD8 T cells, and reducing tumor-infiltrating regulatory T cells (Tregs). Overall, our data show that the effect of TNF blockade on immune responses and clinical outcomes varies depending on the type of TNFi. Certolizumab improves the efficacy of ICI therapy, while infliximab is better at improving tolerability. Therefore, the choice of TNFi to combine with ICI therapy depends on medical need.

## Results

### Different clinical activity in melanoma patients treated with nivolumab and ipilimumab in combination with certolizumab or infliximab

#### Patient characteristics

Thirty-two patients with advanced melanoma (stage IIIc/IV) were treated with a combination of nivolumab and ipilimumab alongside either certolizumab (n=20) or infliximab (n=12) (Extended data Fig. 1a). The demographic and baseline characteristics of the patients are shown in Table 1. Stage IV disease was present in 15 patients (75%) in the certolizumab cohort and 7 patients (58.3%) in the infliximab cohort. Among them, five patients (33.3%) in the certolizumab cohort and one patient (14.3%) in the infliximab cohort presented liver metastases at screening. Overall, 60% and 25% of patients in the certolizumab and infliximab cohorts, respectively, had a BRAF V600 mutation. The median number of injections of TNFi was lower in the certolizumab cohort (2, range [1-14]) than in the infliximab cohort (3, range [1-8]); with 13 (65%) and 4 (33.3%) patients, discontinuing treatment due to toxicity, respectively. Conversely, 4 (20%) and 6 (50%) patients in the certolizumab and infliximab cohorts, respectively, discontinued treatment due to disease progression or death (Extended data table 1).

**Table 1:** TICIMEL patients’ demographic and baseline characteristics.

|  | **Certolizumab**<br>**(N=20)**<br>**N (%)** | **Infliximab**<br>**(N=12)**<br>**N (%)** |
| --- | --- | --- |
| **Sex (N=32)** |  |  |
| Male | 13 (65) | 8 (66.7) |
| Female | 7 (35) | 4 (33.3) |
| **Age at inclusion (N=32)** |  |  |
| Median | 55.5 | 60 |
| Range | (33:73) | (28:75) |
| **ECOG Performance status (N=32)** |  |  |
| OMS 0 | 15 (75) | 10 (83.3) |
| OMS 1 | 4 (20) | 2 (16.7) |
| OMS 2 | 1 (5) | 0 (0) |
| **LDH (U/L) (N=32)** |  |  |
| Normal | 7 (35) | 5 (41.7) |
| ANCS | 11 (55) | 7 (58.3) |
| ACS | 2 (10) | 0 (0) |
| **AJCC stage at screening (N=32)** |  |  |
| Stage IIIc | 5 (25) | 5 (41.7) |
| Stage IV | 15 (75) | 7 (58.3) |
| **If stage IV at screening, number of metastatic sites (N=22)** |  |  |
| 1 site | 5 (33.3) | 1 (14.3%) |
| 2 sites | 5 (33.3) | 4 (57.1%) |
| 3 sites | 2 (13.3) | 2 (28.6%) |
| 5 sites | 1 (6.7) | 0 (0) |
| 6 sites | 1 (6.7) | 0 (0) |
| 8 sites | 1 (6.7) | 0 (0) |
| **If stage IV at screening, liver metastasis (N=22)** |  |  |
| No | 10 (66.7) | 6 (85.7) |
| Yes | 5 (33.3) | 1 (14.3) |
| **BRAF V600 (N=32)** |  |  |
| Unmutated | 8 (40) | 9 (75) |
| Mutated | 12 (60) | 3 (25) |
| **NRAS (N=31)** |  |  |
| Unmutated | 18 (90) | 9 (81.8) |
| Mutated | 2 (10) | 2 (18.2) |
| Missing | 0 | 1 |
| **Prior adjuvant therapy (N=32)** |  |  |
| No | 18 (90) | 11 (91.7) |
| Yes* | 2 (10) | 1 (8.3) |

Abbreviations: AJCC, American Joint Committee on Cancer; ANCS, abnormal not clinically significant; ACS, Abnormal clinically significant; LDH, lactate dehydrogenase.
\*Infliximab cohort: one patient received interferon alpha ten years prior to inclusion
Certolizumab cohort: two patients received anti-PD-1 (nivolumab) therapy prior to inclusion

#### Tolerability and anti-tumor activity

Grade 3/4 drug-related adverse events were observed in 15 (75%) and 4 (33.3%) patients in the certolizumab and infliximab cohorts, respectively (Fig. 1a and Extended data table 2). The most common serious adverse events in the certolizumab cohort were related to the digestive tract (e.g., hepatobiliary and gastrointestinal disorders). Notably, no cases of colitis were reported in the infliximab cohort, whereas 3 (15%) patients in the certolizumab cohort developed grade 2-3 colitis. In addition, skin hypopigmentation (e.g., vitiligo), a feature associated with favorable outcomes^13^, was observed in 5 (25%) and 1 (8.3%) patients of the certolizumab and infliximab cohorts, respectively (Fig. 1b). Anti-tumor activity is shown in Figures 1c-d and Extended data Figs. 1b-c. Importantly, 7 out of 20 (35%) patients treated with ICI and certolizumab and 2 out of 12 (16.7%) patients treated with ICI and infliximab achieved complete responses (Fig 1e).

**Figure 1.**
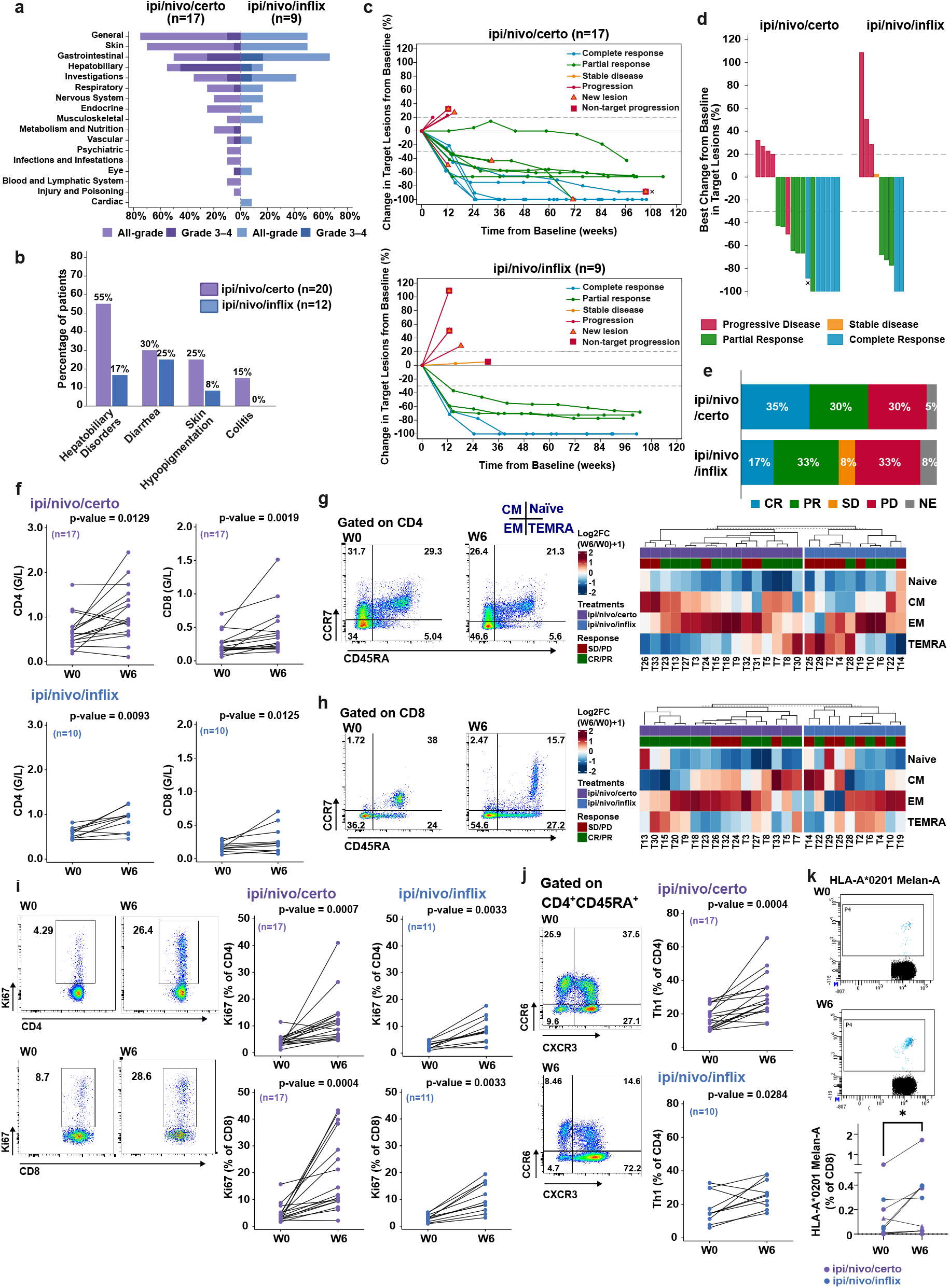
Different clinical activity in patients treated with ICI in combination with certolizumab or infliximab. **a**, Proportion of patients exhibiting treatment-related adverse events (TRAEs) organized by system organ in patients treated with nivolumab and ipilimumab combined with certolizumab or infliximab. The grade of TRAEs is color coded. **b**, Bar graph showing the direct comparison of the percentage of patients experiencing skin hypopigmentation, diarrhea, colitis, and hepatitis in patients treated with ipilimumab, nivolumab and certolizumab or infliximab. **c**, Evolution of metastatic lesions in patients treated with nivolumab and ipilimumab combined with certolizumab (upper panel) or infliximab (lower panel). One patient in complete and one in partial response from the certolizumab and infliximab cohort, respectively, were not evaluable as per RECIST criteria at the first two tumor assessments; one patient from the certolizumab cohort and one from the infliximab cohort progressed before the first tumor assessment; one patient from the certolizumab cohort and one patient from the infliximab cohort died before the first tumor assessment and they are not included in this graph. x: patient in complete response despite 2 persistent lymph node lesions (<10 mm). **d**, Best response in all patients except the same patients mentioned in the legend to “c”. x: patient in complete response despite 2 persistent lymph node lesions (<10 mm). **e**, Graph comparing the proportions of complete responses (CR), partial responses (PR), stable diseases (SD), progressive diseases (PD) and non-evaluable (NE) between patients treated with ipilimumab, nivolumab and certolizumab (ipi/nivo/certo) or infliximab (ipi/nivo/inflix). **f-h**, Blood from patients treated with ipilimumab and nivolumab combined with certolizumab or infliximab was collected before treatment initiation (week 0, W0) and 6 weeks after the beginning of treatments (week 6, W6), and directly assessed for the presence and maturation of T cells by flow cytometry. **f**, Total number of circulating CD4 (left panels) and CD8 (right panels) T cells in the blood of patients treated with ipi/nivo/certo or ipi/nivo/inflix between W0 and W6. **g and h**, Representative staining (left panels) and heatmaps (right panels) showing the evolution between W0 and W6 of proportions of naïve (CCR7^+^CD45RA^+^), central memory (CM, CCR7^+^CD45RA^-^), effector memory (EM, CCR7^-^CD45RA^-^) and effector memory CD45RA^+^ (TEMRA, CCR7^-^CD45RA^+^) CD4 (g) and CD8 (h) T cells in the blood of patients treated with ipi/nivo/certo or ipi/nivo/inflix. **i**, Representative staining (left panels) and proportions (right panels) of proliferating (Ki67^+^) CD4 (upper panels) and CD8 (lower panels) T cells assessed on W0 and W6 PBMC from patients treated with ipi/nivo/certo or ipi/nivo/inflix. **j**, Evolution of proportions of Th1-like CD4 T cells (CXCR3^+^CCR6^-^) between W0 and W6 in the blood of patients treated with ipi/nivo/certo and ipi/nivo/inflix. Right panels, representative staining. k, Proportion of circulating MelanA-specific CD8 T cells between W0 and W6 in HLA-A2 TICIMEL patients. Statistics: Wilcoxon matched-pairs signed-rank test.

#### Pharmacodynamic assessment of systemic T cell responses

Analyses of the effect of combining ICI with certolizumab or infliximab on circulating T cell responses between baseline (week 0, W0) and week 6 (W6) showed similar increases in the numbers and proportions of proliferating CD4 and CD8 T cells and maturation towards central memory and effector memory phenotypes (Figs. 1f-i). Increases in the proportions and numbers of circulating Th1-like cells (CD4^+^CXCR3^+^CCR6^-^) were also observed in both cohorts (Fig. 1j). No variations were observed for Th2, Th17 and Th17-Th1-like CD4 T cells (Extended data Fig. 1d). Importantly, these were concomitant to increases in the proportions of circulating MelanA-specific CD8 T cells in patients from both cohorts (Fig. 1k).

Overall, our data suggest that certolizumab and infliximab have different clinical effects when used in combination with nivolumab and ipilimumab in patients with advanced melanoma, although both tritherapies promote the differentiation and proliferation of circulating T cells.

### Infliximab and certolizumab, when combined with ICI, differentially affect the molecular signature of circulating effector T cells

As patients treated with both tritherapies showed different profiles with regard to toxicity and efficacy, we hypothesized that the observed similar modulation of systemic T cell responses (Figs. 1f-k) was only apparent. To evaluate the molecular impact of the two tritherapies more in depth, we performed cellular indexing of transcriptomes and epitopes by sequencing (CITE-Seq) analyses on peripheral blood mononuclear cells (PBMC) from 8 TICIMEL patients per cohort, taken at W0 and W6. We compared these data with those from another cohort of 8 advanced melanoma patients treated with bitherapy (nivolumab and ipilimumab) in the MELANFα clinical trial [NCT03348891])^12^ (Fig. 2a). We then further evaluated the molecular impact of treatments on T cells. Our initial focus was on CD8^+^ T cells, as these play a key role in the clinical outcome of patients treated with ICI.

**Figure 2.**
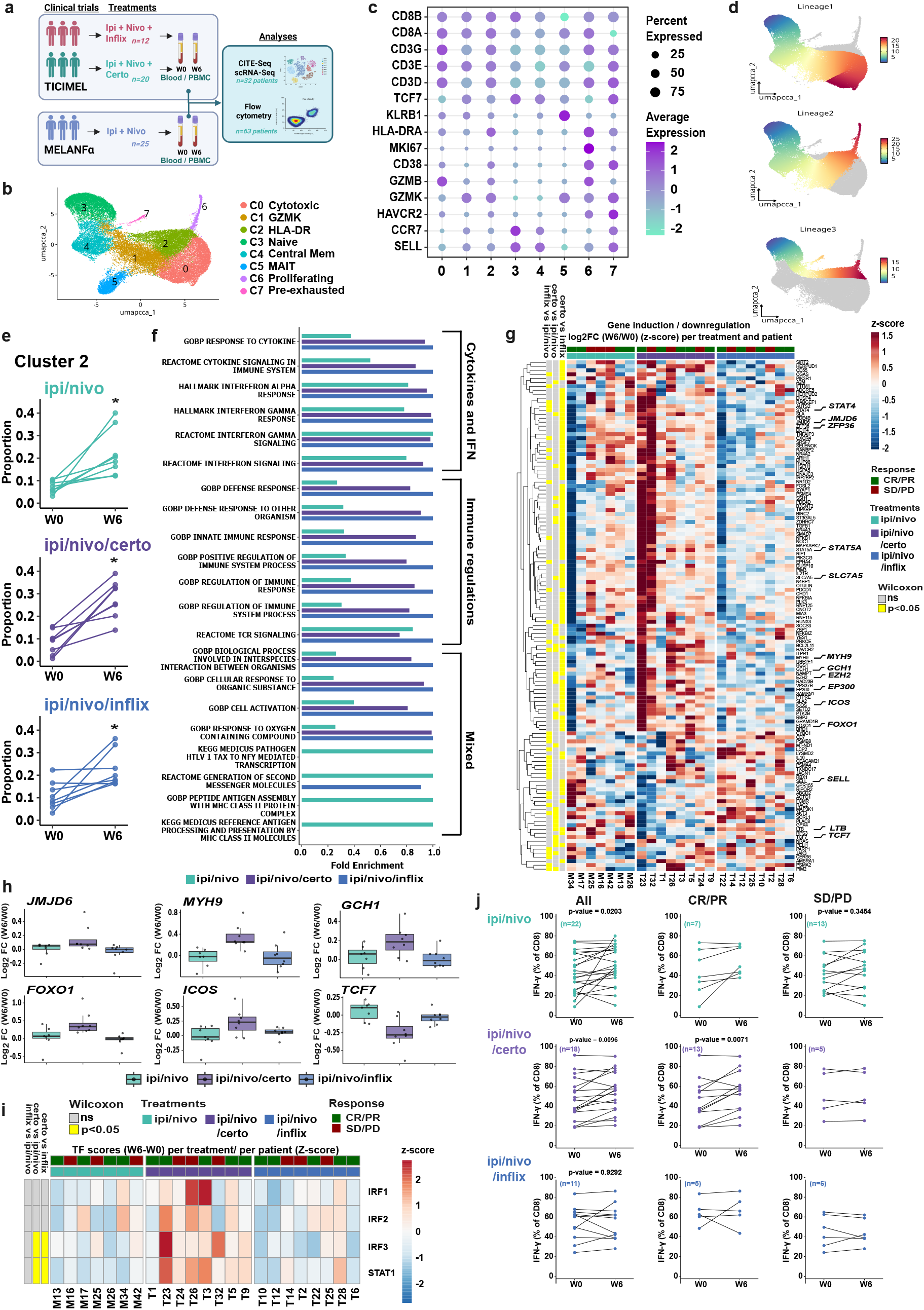
Infliximab inhibits the induction of a molecular program related to T cell activation in circulating effector CD8 T cells during ICI therapy, whereas certolizumab promotes it. **a**, Diagram summarizing the analysis strategy. The phenotype of circulating immune cells of TICIMEL patients was compared to the one of patients treated with nivolumab and ipilimumab alone (MELANFα clinical trial). T cell responses in the blood of patients of all three cohorts (ipi/nivo; ipi/nivo/certo and ipi/nivo/inflix) were analysed at W0 and W6 by CITE-Sequencing (CITE-Seq) and flow cytometry. **b-i**, Peripheral blood monuclear cells isolated from the blood of patients (8 patients per cohort) before treatment initiation (week 0, W0) and 6 weeks after the beginning of treatments (week 6, W6) was anlayzed by CITE-Seq. **b**, Unified Manifold and Approximation and Projection (UMAP) of CD8 T cells of all patients and time points, identifying 8 clusters of CD8 T cells [7 clusters [C0-C7] of CD8 T cells and 1 cluster of Mucosal Associated Invariant T cells (MAIT)]. **c**, Dotplot showing the expression of key gene markers in the identified CD8 subclusters (proportion and average expression). **d**, Trajectory analysis using slingshot and showing 3 possible endpoints. **e**, Proportion of cells from cluster 2 between W0 and W6 per treatment cohort. *p<0.05 Wilcoxon. **f**, A differential gene expression analysis was performed to compare W6 and W0 cells from cluster 2, separating each treatment cohort. Over-representation analysis (ORA) was then performed on the 3 lists of differentially expressed genes (DEG) to identify the 10 most significantly deregulated pathways after ipi/nivo, ipi/nivo/certo or ipi/nivo/inflix (see Extended data Fig. 2). The bar plot shows the fold enrichment of the 21 identified pathways in cells from cluster 2 after ipi/nivo; ipi/nivo/certo or ipi/nivo/inflix. **g**, Genes contributing to the identification of pathways depicted in “f” were identified. Those, which were differentially modulated (log2 fold change (W6/W0) of the gene expression) between at least two treatment cohorts were selected and plotted on a heatmap. Gene induction or downregulation is shown per patient and treatment cohort. On the left side, yellow squares identified a significant difference in gene expression comparing two treatment cohort at a time (p<0.05, Wilcoxon). **h**, Boxplot showing the log2 fold change (W6/W0) in the expression of genes from the heatmap in “g” and involved in T cell activation and metabolism (*JMJD6, MYH9, GCH1*), and differentiation (*FOXO1, ICOS, TCF7*) per treatment cohort. **i**, Heatmap showing the modulation in the activity of IRF1, IRF2, IRF3 and STAT1 transcription factors (TF) in cluster 2 cells between W0 and W6 per patient and treatment cohort (DecouplR package). On the left side, yellow squares identified a significant difference between the evolution of TF activities comparing two treatment cohorts at a time (p<0.05, Wilcoxon; ns: non-significant). **j**, CD8 T cells were isolated from W0 and W6 PBMC of patients before polyclonal restimulation in the presence of vesicular transport inhibitors. IFN-γ production was assessed by flow cytometry. CR: complete response; PR: partial response; SD: stable disease; PD: progressive disease reflecting the best response over the two years of follow-up for TICIMEL patients and response at 12 weeks for MELANFα patients. Statistics: the Wilcoxon matched-pairs signed-rank test was applied to data sets with n>10. For CITE-Seq analyses, one patient from the ipi/nivo cohort was excluded from the analyses in Fig. 2g-i because fewer than 10 cells were detected in cluster 2 at one time point.

Total PBMC single cells from all patients and time points were integrated and analyzed together. After clustering and dimensional reduction using Uniform Manifold Approximation and Projection (UMAP), CD8 T cells were isolated and analyzed independently. Non-supervised clustering divided CD8 T cells into eight clusters (Fig. 2b). These clusters were identified on their specific gene (Fig 2c) and protein expression (Extended data Fig. 2a). Trajectory analysis revealed that naïve T cells (cluster 3, expressing the *SELL, CCR7*, and *TCF7* genes, and the CD62L, CD45RA proteins) differentiated into three subtypes: (i) cytotoxic T cells (cluster 0, expressing *GZMB*); (ii) proliferating T cells (cluster 6, expressing *MKI67*); and (iii) HLA-DR-positive cells (cluster 2, expressing *HLADRA*) (Fig. 2c, Extended Data Fig. 2a). All treatments significantly increased the proportion of cells in clusters 2 (C2) and 6 (C6) (Fig 2e and extended data Fig. 2b). However, as there were insufficient cells in C6 to assess the molecular impact of the treatments, we focused on cells in C2, expressing *HLADRA*. Then, we identified differentially expressed genes (DEG), followed by over-representation analyses (ORA) of biological pathways comparing W6 to W0 cells from C2 according to treatment. A strong overlap was observed on the significantly dysregulated pathways across all treatments. The 10 most significantly dysregulated pathways in each cohort (Extended data Figs. 2c-e) were selected and their enrichment in all treatment groups was calculated. Pathways related to “cytokines or IFN signaling” and “immune regulation” were enriched in C2 cells after ICI and certolizumab or infliximab treatment in contrast to ICI treatment alone (Fig. 2f). The genes contributing to the selection of these pathways revealed that 40% exhibited significant differential modulation depending on treatments (Fig. 2g). Most strikingly, the expression of genes involved in T cell activation (*RNF125, MYH9, JMJD6*, and *ZFP36*), differentiation (*ICOS, STAT4, FOXO1*, and *EZH2*) and metabolic fitness (*EP300, GCH1*, and *SLC7A5*), was increased following the tritherapy with certolizumab. However, the combination of ICI and infliximab did not elicit this response. C2 cells from patients treated with ICI alone tended to exhibit an intermediate phenotype (Figs. 2g and h). C2 cells from patients treated with ICI and certolizumab also showed a decrease in the expression of genes related to the naïve state (*TCF7, SELL* and *LTB*) (Figs. 2g and h). Finally, we observed an increase in the activity of transcription factors related to IFN signaling (IRF1/2/3, STAT1) in C2 cells from patients treated with ICI and certolizumab, compared to those receiving ICI alone or in combination with infliximab (Fig. 2i).

Next, we evaluated the ability of CD8 T cells, isolated from PBMC, to produce IFN-γ after polyclonal activation. We found a significant increase of IFN-γ-producing CD8 T cell proportions in patients from the bitherapy and certolizumab cohorts, but not for those from the infliximab cohort. Furthermore, this increase was significantly associated with a better response to treatment with ICI and certolizumab, and a similar tendency was observed in patients treated with ICI alone (Fig. 2j).

Lastly, we also assessed how treatments impacted the molecular profiles of CD4 T cells. We identified 11 clusters including two naïve clusters (expressing the *TCF7* and *CCR7* genes, and CD62L, CD45RA proteins), one central memory cluster (expressing the *TCF7* and *CCR7* genes, but not the CD45RA protein), three clusters of activated effector cells and one cluster of exhausted CD4 T cells (Extended data Fig. 3a-d). Among those, cluster C1, which exhibited an early activated phenotype (*TCF7, KLRB1, ICOS*) and intermediate levels of Th1 (*TBX21, CXCR3*), Th2 (*GATA3, CCR4*) and Th17 (*RORA, CCR6*) markers, increased following bi- and tritherapy (Extended data Figs. 3b, c and e). Using the strategy as described above for CD8 T cells, we found 18 significantly deregulated pathways in this cluster between W6 and W0 after bi-or tritherapy (Extended data Fig 3f). These pathways were related to IFN signaling and activation of immune responses, with 24.6% of genes contributing to these pathways being significantly deregulated between at least two treatment cohorts (Extended data Fig. 3g). Some of these genes were upregulated after ICI and certolizumab, but not infliximab, and others were involved in the Th1/Th17 differentiation and activation (*ARID5A, SKI, ZEB1* and *IRS2*). Finally, after certolizumab and ICI, the estimated activity of the transcription factors STAT1, IRF1/2/3 and T-bet, also tended to increase in these cells (Extended data Fig. 3h).

**Figure 3.**
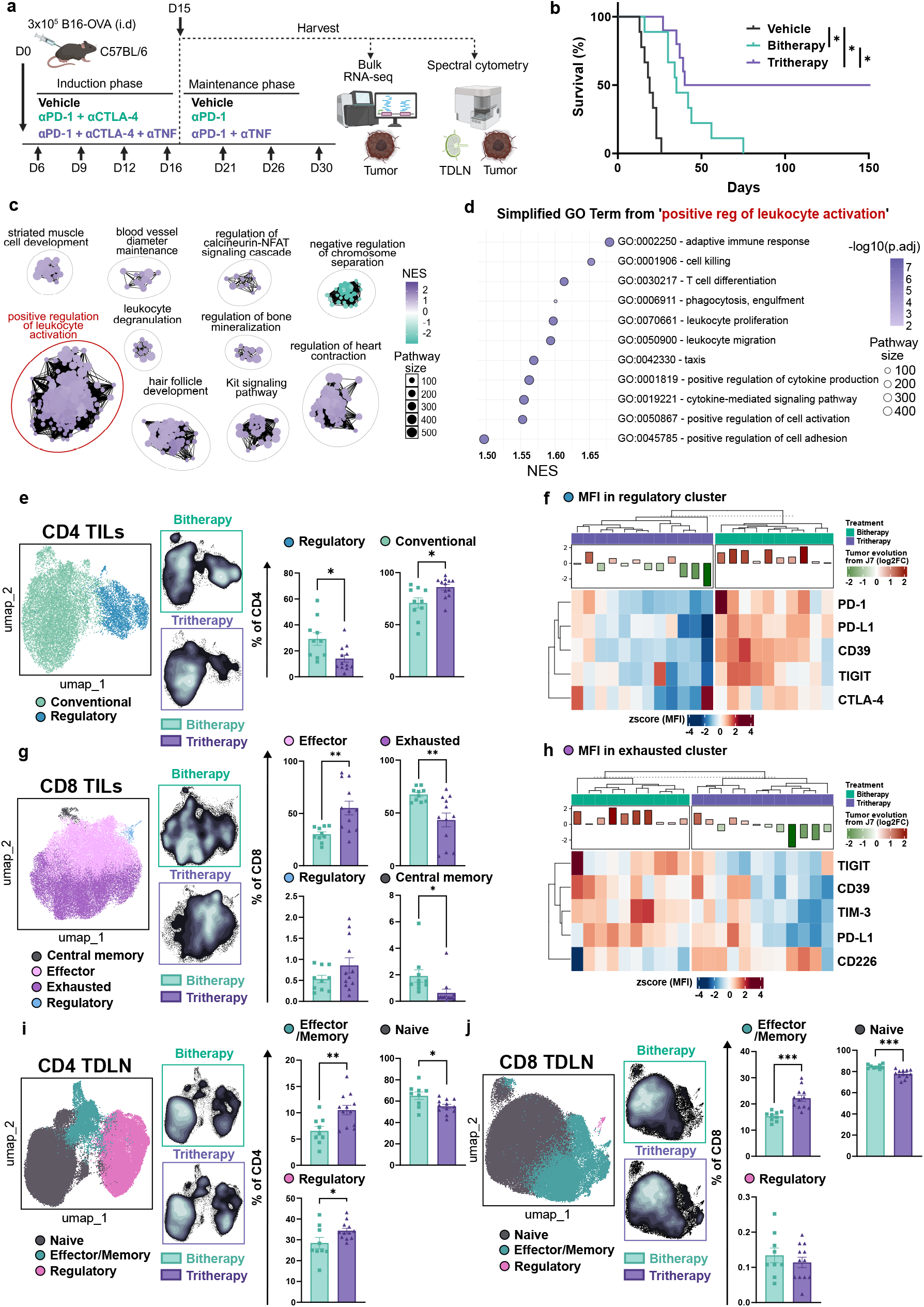
TNF blockade can improve the immune response and efficacy of anti-PD-1 and anti-CTLA-4 in mice with melanoma. **a**, Schematic representation of the murine therapeutic regimen. Wild-type C57BL/6 mice were injected intradermally with B16-OVA cells. Starting on day 6, mice received four induction cycles of vehicle, anti–PD-1 plus anti–CTLA-4 (bitherapy), or bitherapy combined with anti-TNF (tritherapy), followed by three maintenance cycles of vehicle, anti–PD-1 alone, or anti–PD-1 plus anti-TNF. In a separate set of experiments, tumors and tumor-draining lymph nodes (TDLN) were collected on day 17 for bulk RNA-Seq or spectral cytometry analysis. **b**, Kaplan–Meier survival curves of mice according to treatment (n=9-10 mice per group). **c**, aPEAR enrichment network analysis comparing tritherapy versus bitherapy (n=6 mice per group) based on DEGs in tumor bulk RNA-Seq. **d**, Simplification of the cluster “positive regulation of lymphocyte activation” (highlighted in red in (c)), using rrvgo package and showing enriched GO terms. **e**, UMAP projection of CD4^+^ tumor-infiltrating lymphocytes (TILs), identifying conventional and regulatory subsets, with UMAP plots stratified by treatment and histograms showing the frequency of each subset (n=10-12 mice per group). **f**, Heatmap of the median fluorescence intensity (MFI) of differentially expressed markers on Tregs between groups, with an associated waterfall plot showing log_2_ fold change in tumor volume between day 6 and day 17. **g**, UMAP projection of CD8^+^ TILs (left) identifying four clusters—central memory, effector, exhausted, and regulatory—with treatment-specific UMAPs and histograms of cluster frequencies (right) (n=10-12 mice per group). **h**, Heatmap of the MFI of differentially expressed markers on the exhausted CD8^+^ T cell cluster. **i**, UMAP projection of CD4^+^ TDLN cells identifying naïve, effector/memory, and regulatory subsets, with histograms showing their frequency across treatments. **j**, UMAP projection of CD8^+^ TDLN subsets identifying naïve, effector/memory, and regulatory subsets, with histograms showing their frequency across treatments. Statistics: survival curves were compared using the log-rank (Mantel–Cox) test. For other comparisons, data were analyzed using unpaired two-tailed t tests. P values <0.05 were considered significant (*P<0.05; **P<0.01; ***P<0.001; ****P<0.0001).

As TNF has been shown to promote the survival and effector properties of Tregs^14-17^, we then assessed the impact of the two tritherapies on circulating Tregs in patients. Circulating proportions and absolute numbers of Tregs were increased in all cohorts between W0 and W6 (Extended data Fig. 4a). In patients treated with ICI alone, identification of DEG between W0 and W6 Tregs showed an increased expression of genes related to the regulation of Treg function and response to IFN signaling (*HDAC9, TOX2, STAT1*), mitochondrial metabolism (*PCCA, CBR4*) or cell migration (*MIEN1, ABI1*). However, this was less pronounced when ICI therapy was combined with certolizumab or infliximab (Extended data Fig. 4b and c). In patients treated with ICI alone, ORA analyses on DEG further showed an increased expression of pathways related to IFN-γ signaling and immune activation (Extended data Fig. 4d).

Overall, our results show that infliximab decreases the expression of genes related to CD8 T cell activation induced by ICI and, to a lesser extent, in CD4 effector T cells, whereas certolizumab tends to increase it. Certolizumab also increases the expression of a gene signature related to IFN sensing in effector CD8 T cells. This is associated with increased proportions of circulating CD8 T cells that can produce IFN-γ in good responders. Finally, both certolizumab and infliximab inhibit the ICI-mediated induction of a molecular program related to cell activation, migration and IFN sensing in Tregs.

### TNF blockade can enhance the immune response and the efficacy of the combination of anti-PD-1 and anti-CTLA-4 in mouse melanoma models

To further evaluate the effect of TNF blockade in combination with ICI, we conducted experiments using preclinical mouse melanoma models. We designed a treatment regimen mimicking the one given to patients, and comprising an induction phase (anti-PD-1 + anti-CTLA-4 ± anti-TNF) followed by a maintenance phase (anti-PD-1 ± anti-TNF) (Fig. 3a). Compared to the vehicle control group, the survival of mice treated with either bi-or tritherapy was longer (Fig. 3b). While mice treated with bitherapy showed partial but not complete responses, 50% of mice treated with tritherapy had complete responses (Fig. 3b and Extended Data Fig. 5a). Rechallenging mice that achieved complete responses with B16-OVA cells in the absence of treatment led to a complete tumor cell rejection in 3 out of 5 mice. Only 2 mice developed slow-growing tumors, indicating the establishment of long-term immune protection following our tritherapy regimen (Extended Data Fig. 5b). Next, we investigated at which phase of treatment the anti-TNF enhanced ICI efficacy, by administering anti-TNF during the induction or maintenance phase. Alternatively, mice were treated with anti-TNF and ICI in both phases. TNF blockade enhanced efficacy when delivered during the induction phase or both phases, but not when delivered during the maintenance phase alone, indicating a role in priming anti-tumor responses (Extended Data Fig. 5c).

To gain insight into how TNF blockade enhances the efficacy of ICI during the induction phase, we performed bulk RNA Sequencing (RNA-Seq) on B16-OVA tumors from mice treated with bi-or tritherapy (Fig. 3c). Gene set enrichment network analysis (GSEA) using aPEAR package revealed the enrichment of a cluster of more than 200 pathways associated to “positive regulation of leukocyte activation” in the tritherapy group. Then, by identifying the redundance of terms (rvvgo package), we related these pathways to “adaptive immune response”, “cell killing”, and “T cell differentiation” processes (Figs. 3c and d). Accordingly, tumors from tritherapy-treated mice displayed higher expression levels of *Prf1, Gzmb, Klrk1, Ifng* and *Tbx21*, which are all related to T cell activation and cytotoxicity (Extended Data Fig. 5d).

To further evaluate the pharmacodynamic effects of anti-TNF in combination with ICI, we used spectral flow cytometry to compare the impact of bi- and tritherapies on tumor-infiltrating lymphocytes (TILs) and tumor-draining lymph nodes (TDLNs) at the end of the induction phase. The proportion of tumor-infiltrating conventional CD4^+^ T cells was significantly higher after tritherapy than after bitherapy, while the proportion of Tregs was significantly lower (Fig. 3e and Extended data Fig. 5e). Furthermore, the proportion of CD39^+^ TIGIT^+^ Tregs was significantly lower after tritherapy (Extended Data Fig. 5f) as were the expression levels of CD39, CTLA-4, PD-1, TIGIT, and PD-L1 (Fig. 3f), suggesting a decrease in their immunosuppressive phenotype.

Regarding CD8^+^ TILs, the proportion of effector T cells (i.e., early activated) was higher upon tritherapy, while that of exhausted T cells (PD-1^+^TIM-3^+^CD39^high^), central memory T cells (CD62L^+^CD44^+^SLAMF6^+^) and dead cells was lower (Fig. 3g and Extended Data 5g and h). The median fluorescence intensity (MFI) of the immunosuppressive molecules TIGIT, CD39, TIM-3 and PD-L1 was significantly lower on exhausted CD8 TILs after tritherapy (Fig. 3h). The proportion of total PD-1^+^TIM-3^+^CD39^+^ CD8^+^ T cells was lower in tumors of tritherapy-treated mice than in those treated with bitherapy (Extended data Fig. 5i). In contrast, the MFI of CD226, a positive co-stimulatory molecule^18,19^, as well as the proportion of cells expressing high levels of CD226 was significantly higher upon tritherapy (Fig. 3h and Extended data Fig. 5j). These results suggest that the tritherapy increases the activation and decreases the exhaustion of CD8^+^ TILs compared to the bitherapy. In TDLNs, the tritherapy decreased the proportion of naïve T cells while increasing that of effector CD4^+^ and CD8^+^ subsets (Figs. 3i and j and Extended data Figs. 5k and l). The proportion of CD4^+^ Tregs was also slightly, yet significantly, higher with tritherapy, while the proportion of CD8^+^ Tregs was marginal and did not differ significantly between the two therapies (Figs. 3i and j).

We also investigated the putative benefits of the tritherapy in the poorly immunogenic B16-F10 model (Extended data Fig. 5m). We observed delayed tumor growth in some mice treated with tritherapy compared to those treated with bitherapy (Extended data Fig. 5n). However, neither regimen induced complete responses. As in the B16-OVA model, we observed reduced Treg infiltration in B16-F10 tumors in tritherapy-treated mice compared to the control and bitherapy groups (Extended Data Fig. 5o).

Taken together, our preclinical data suggest that anti-TNF antibodies do not interfere with, and even enhance, the efficacy of anti-PD-1 and anti-CTLA-4, by limiting the infiltration of Tregs and the exhaustion of CD8^+^ T cells in tumors.

### Certolizumab, but not infliximab, enhances the efficacy of anti-PD-1 and anti-CTLA-4 in TNF- and TNFR2-humanized mice bearing melanoma

A major limitation of preclinical mouse models is that most TNFi that target human TNF, such as certolizumab and infliximab, do not interact with murine TNF. Therefore, we used mice humanized for TNF and TNFR2 (hTNFKI×hTNFR2KI)^17^, which carry a functional human TNF signaling module, with the human TNF being able to activate mouse TNFR1^20^. We hence studied whether certolizumab and infliximab potentiate ICI efficacy in mice bearing B16-OVA tumors, using a treatment regimen similar to that used for TICIMEL patients (Figure 4a). Consistent with our observations in patients, certolizumab significantly improved ICI efficacy and overall survival (Figs. 4b and c). No improvement was observed with infliximab. This result was not restricted to B16-OVA, certolizumab but not infliximab, also enhanced the effectiveness of ICI therapy in the Yumm1.7 mouse melanoma model (Extended data Figs. 6a-c).

**Figure 4.**
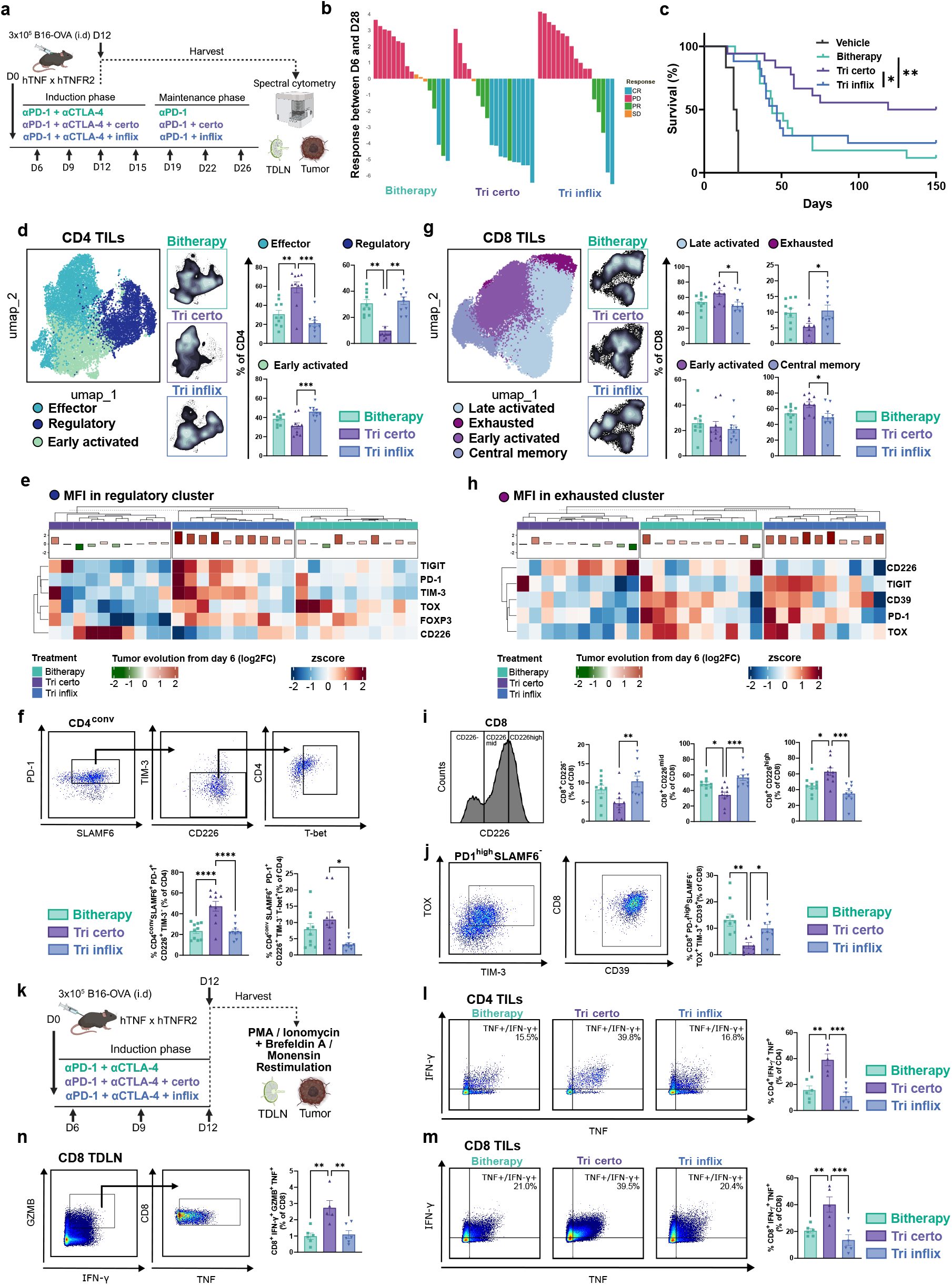
Certolizumab, but not infliximab, enhances the efficacy of anti-PD-1 and anti-CTLA-4 combination in TNF and TNFR2-humanized mice. **a**, Schematic representation of the murine therapeutic regimen. **b**, Waterfall plot showing log_2_ fold change in tumor volume between day 6 and day 28 for each treatment group (n= 17 to 18 mice per group). **c**, Kaplan– Meier survival curves of mice according to treatment. **d**, Left, UMAP projection of CD4^+^ tumor-infiltrating lymphocytes (TILs), identifying three clusters: effector, regulatory (Tregs), and early activated subsets. Right, UMAP plots stratified by treatment with corresponding histograms showing the frequency of each CD4^+^ T cell cluster (n=10 mice per group). **e**, Heatmap of the MFI of differentially expressed markers in regulatory CD4^+^ T cells cluster across the three treatment groups, together with a waterfall plot showing log_2_ fold change in tumor volume between day 6 and day 12. **f**, Manual gating strategy (top) and frequency (bottom) of activated conventional CD4^+^ T cells (SLAMF6^+^PD-1^+^CD226^+^TIM-3^−^), with the proportion of T-bet^+^ Th1 cells among them shown on the right. **g**, Left, UMAP projection of CD8^+^ TILs identifying four clusters: early activated, exhausted, late activated, and central memory subsets. Right, UMAP plots stratified by treatment with histograms showing the frequency of each CD8^+^ T cell cluster. **h**, Heatmap of the MFI of differentially expressed markers among exhausted CD8^+^ T cells. **i**, Expression levels of CD226 in CD8^+^ TILs categorized as negative, mid, or high, with representative gating (left) and frequencies (right). **j**, Gating strategy (left) and histogram (right) showing the frequency of exhausted CD8^+^ TILs (PD-1^high^SLAMF6^−^TOX^+^TIM-3^+^CD39^+^). **k**, Schematic diagram showing the cytokine production assay. **l**, Representative plots (left) and quantification (right) of TNF and IFN-γ production by CD4^+^ TILs (n=5-6 mice per group). **m**, Same as in (l), but for CD8^+^ TILs. **n**, Gating strategy (left) and quantification (right) of CD8^+^ T cells from TDLN producing IFN-γ, TNF, and granzyme B. Statistics: survival curves were compared using the log-rank (Mantel–Cox) test. For other comparisons, data were tested for normality and analyzed using one-way ANOVA with appropriate post hoc correction or Kruskal–Wallis test. P values <0.05 were considered significant (P<0.05; *P<0.01; **P<0.001; ***P<0.0001).

After two treatment cycles, we performed the immune profiling of B16-OVA tumors using spectral cytometry, focusing on CD4 and CD8 TILs. Higher proportions of effector CD4^+^ T cells and lower proportions of Tregs were observed in tumors of mice treated with the certolizumab tritherapy compared to the bitherapy and infliximab tritherapy groups (Fig 4d, Extended data Figs. 6d and e). Within Tregs population, the expression level of TOX, PD-1, TIM-3, TIGIT and FOXP3 was reduced in the certolizumab tritherapy group, while that of CD226 was increased (Fig. 4e). Conversely, tumor-infiltrating Tregs in mice treated with the infliximab tritherapy expressed higher levels of TIM3 and FOXP3 and were more viable (Fig 4e and Extended data Fig. 6f). This may reflect a higher immunosuppressive phenotype^21-23^. The proportion of activated cells (PD1^+^TIM-3^-^CD226^+^) and activated Th1 (T-bet^+^) was higher in mice treated with the certolizumab tritherapy (Fig. 4f). Moreover, a low proportion of cell death was observed in conventional CD4^+^ TILs, and this was significantly reduced in the certolizumab tritherapy treated mice (Extended data Fig. 6g).

Similar effects were observed in CD8^+^ TILs, with higher proportion of central memory and late activated effector cells, and lower proportion of exhausted cells in mice receiving the certolizumab tritherapy than in those receiving bitherapy or the infliximab tritherapy (Fig. 4g). This was associated with decreased levels of TOX, PD-1, CD39 and TIGIT and increased levels of CD226 on exhausted CD8^+^ TILs in the certolizumab tritherapy group (Fig. 4h). Consequently, the proportion of cells expressing high levels of CD226 was significantly higher in mice treated with the certolizumab tritherapy group (Fig. 4i). Moreover, the proportion of exhausted cells (PD-1^high^ SLAMF6^-^ TOX^+^ TIM-3^+^ CD39^+^) and dead cells was lower in the certolizumab tritherapy group (Figs. 4j and Extended Data Fig. 6h).

We then assessed the capacity of CD4^+^ and CD8^+^ TILs to produce IFN-γ after polyclonal restimulation (Fig. 4k). Interestingly, the decreased exhaustion of TILs observed after treatment with the certolizumab tritherapy was accompanied by a higher proportion of CD4^+^ and CD8^+^ TILs able to produce TNF and IFN-γ, when compared to TILs from bitherapy or infliximab tritherapy treated mice (Figs. 4l and m).

In the TDLNs, two rounds of certolizumab tritherapy led to increased proportions of both effector CD4^+^ and CD8^+^ T cells (IFN-γ^+^TNF^+^GZMB^+^), as well as of Th1 cells (Fig. 4n and Extended data Figs. 6i-n). Of note, the proportion of Th1 cells was also increased upon infliximab tritherapy compared to bitherapy (Extended data Fig. 6k). The certolizumab tritherapy also favored a higher proliferation of activated CD8^+^ T cells (PD-1^+^Ki67^+^) and an increased proportion of CD8^+^ precursor exhausted T cells (CD8^+^ Tpex) (TCF1^+^SLAMF6^+^) compared to bitherapy (Extended data Figs. 6n and o). A tendency to increase was also observed for CD8^+^ Tpex in mice treated with the infliximab tritherapy. Importantly, higher proportions of cytotoxic CD8 T cells capable of producing Granzyme B, TNF and IFN-γ were detected in TDLN of mice treated with certolizumab tritherapy compared to those treated with bitherapy or infliximab tritherapy (Fig 4n).

Overall, these preclinical data show that, unlike infliximab, certolizumab reduces T cell exhaustion and Tregs proportion in tumors, and promotes T cell activation in both tumors and TDLNs when combined with ICI.

### The infliximab Fc domain reduces the effectiveness of TNF blockade when used in combination with ICI

Since infliximab contains an Fc fragment that can affect immune responses and immunogenicity in mice, while certolizumab does not, we replaced the human IgG1 Fc portion of infliximab with either a mouse IgG1 (functionally equivalent to human IgG4)^24^ or a mouse IgG2a (functionally equivalent to human IgG1)^25^ Fc fragment (Fig. 5a) in order to evaluate their impact. Mice injected with B16-OVA melanoma cells were treated with either tritherapies containing certolizumab, infliximab, mouse IgG1 infliximab or mouse IgG2a infliximab (Fig. 5b). Tritherapy with certolizumab or mouse IgG1 infliximab, but not human IgG1 or mouse IgG2a infliximab, enhanced the rate of objective responses (i.e., PR and CR) (Fig. 5c). This demonstrates that the infliximab Fc fragment is responsible for counteracting the benefits of TNF blockade. To gain insight into the mechanisms by which mouse IgG1 infliximab and certolizumab enhance the efficacy of ICI, we first performed bulk RNA-Seq on B16-OVA tumors, comparing mice treated with bitherapy with those treated with certolizumab, infliximab, or mouse IgG1 infliximab tritherapy. GSEA analyses of tumors from these mice revealed an enrichment of GO pathways associated with “positive regulation of lymphocyte activation” in groups receiving certolizumab or mouse IgG1 tritherapy, but not in those treated with human IgG1 infliximab tritherapy (Fig 5d). Consistent with this finding, genes encoding proteins involved in T cell activation or cytotoxic activity were highly expressed in tritherapies containing either certolizumab or mouse IgG1 infliximab, compared to bitherapy or the tritherapy containing human IgG1 infliximab (Fig. 5e).

**Figure 5.**
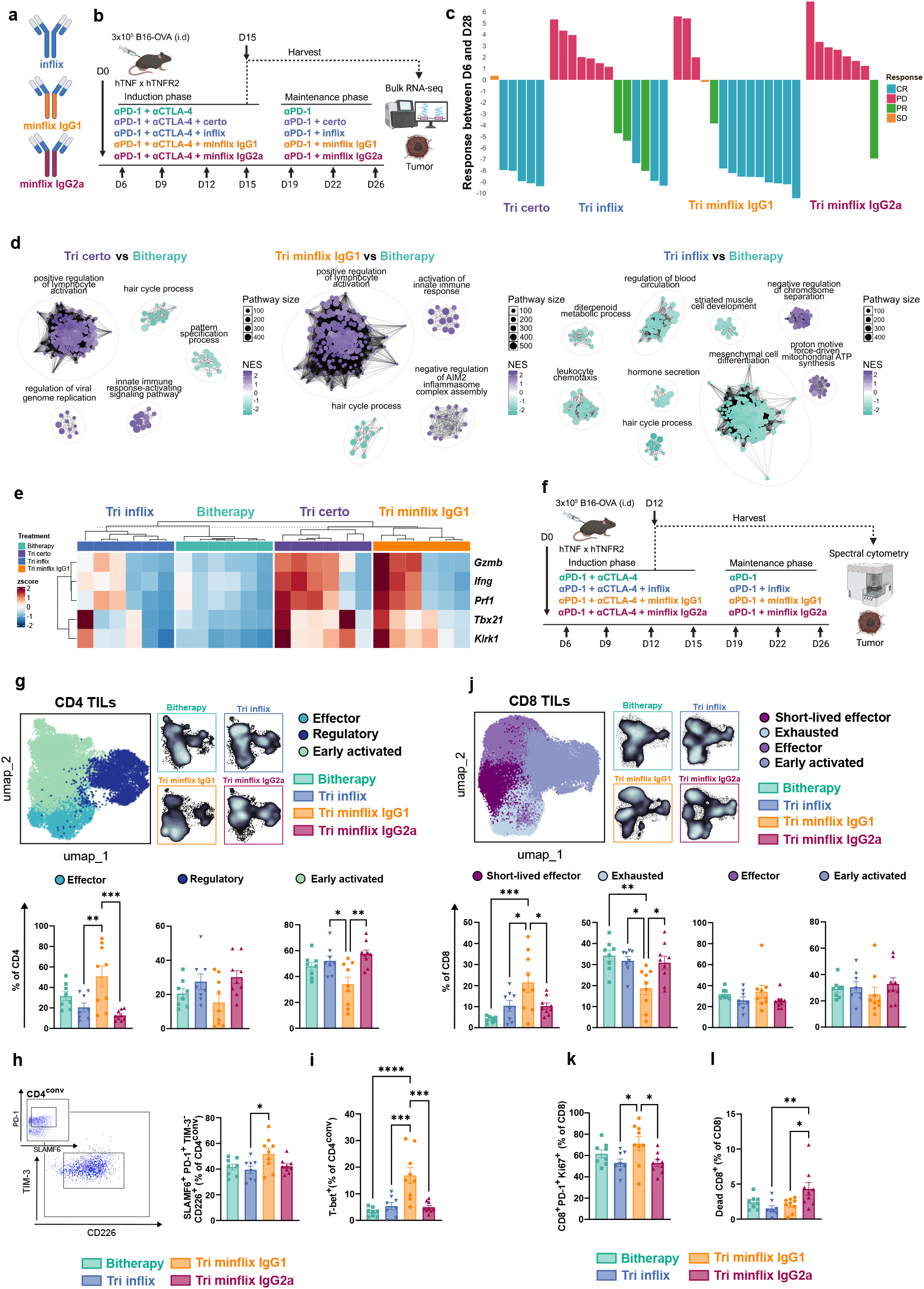
The infliximab Fc fragment reduces the effectiveness of TNF blockade when used in combination with ICI. **a**, Schematic representation of the three infliximab constructs: infliximab (top), infliximab with a murine IgG1 Fc (middle), and infliximab with a murine IgG2a Fc (bottom). **b**, Treatment protocol: hTNFxhTNFR2 mice bearing B16-OVA tumors were treated as in Fig. 4a, with anti–PD-1 plus anti–CTLA-4 alone or in combination with certolizumab, infliximab, mIgG1-Fc infliximab, or mIgG2a-Fc infliximab. On day 15, tumors from mice treated with dual ICI, certolizumab, wild-type infliximab, or mIgG1-Fc infliximab were collected for RNA extraction and bulk RNA-Seq analysis. **c**, Waterfall plot showing log_2_ fold change in tumor volume between day 6 and day 28 across the four treatment groups (n=6-14 mice per group). **d**, aPEAR enrichment network visualization based on DEGs in tumor bulk RNA-Seq: certolizumab vs bitherapy (left), mIgG1-Fc infliximab vs bitherapy (middle), and infliximab vs bitherapy (right) (n=6 mice per group). **e**, Heatmap of cytotoxicity-related genes differentially expressed between the four groups. **f**, Experimental schematic of tumor-infiltrating lymphocyte analysis: mice treated with bitherapy, infliximab, mIgG1-Fc infliximab, or mIgG2a-Fc infliximab were euthanized on day 12, and tumors were analyzed by spectral flow cytometry (n=8-9 mice per group). **g**, Left, UMAP projection of CD4^+^ TILs identifying effector, regulatory (Tregs), and early activated subsets, with UMAP plots stratified by treatment. Right, histograms showing the frequency of each CD4^+^ TIL cluster per treatment. **h**, Manual gating strategy for activated conventional CD4^+^ T cells (left) with associated histograms showing their frequency across treatments (right). **i**, Frequency of CD4^+^ Th1 cells (T-bet^+^) among conventional CD4^+^ T cells. **j**, Left, UMAP projection of CD8^+^ TILs identifying four clusters: short-lived effector, exhausted, effector, and early activated CD8 T cells, with UMAP plots stratified by treatment. Right, histograms showing the frequency of each CD8^+^ T cell cluster per treatment. **k**, Manual gating quantification of proliferating CD8 T cells (Ki67^+^PD-1^+^). **l**, Frequency of dead CD8 T cells. Statistics: data were tested for normality and analyzed using one-way ANOVA with appropriate post hoc correction or Kruskal–Wallis test. P values <0.05 were considered significant (P<0.05; *P<0.01; **P<0.001; ***P<0.0001).

Following two cycles of treatment, we used spectral flow cytometry to evaluate the proportions of CD4^+^ and CD8^+^ TILs (Fig. 5f and Extended data Figs. 7a and b). The proportion of effector CD4^+^ TILs was significantly higher in the mouse IgG1 infliximab group than in the human IgG1 and mouse IgG2a infliximab groups (Fig. 5g). Conversely, the proportion of central memory CD4^+^ TILs was reduced in the mouse IgG1 infliximab group. We also observed a tendency for Tregs proportions to be lower in tumors from the mouse IgG1 infliximab group, although this difference was not statistically significant (Fig. 5g). As previously observed in certolizumab-treated tumors, the proportions of activated CD4^+^ TILs (PD-1^+^SLAMF6^+^TIM-3^-^CD226^+^) and Th1 cells (T-bet^+^) were significantly higher in the mouse IgG1 infliximab group (Figs. 5h and i). Among the CD8^+^ TILs, the proportions of short-lived effector (T-bet^+^) and exhausted CD8^+^ TILs were significantly higher and lower, respectively, in the mouse IgG1 infliximab group compared with the other three groups (Fig. 5j). Finally, while the proportion of activated CD8^+^ TILs (PD-1^+^Ki67^+^) was higher in the mouse IgG1 infliximab group compared to the human IgG1 and the mouse IgG2a infliximab groups, the proportion of dead cells among CD8^+^ TILs was higher in the mouse IgG2a infliximab group compared to the other two groups (Figs. 5k and l).

Taken together, our preclinical data demonstrated that certolizumab and infliximab have different effects when used in combination with anti-PD-1 and anti-CTLA-4 in TNF- and TNFR2-humanized mice. Furthermore, the data showed that the human IgG1 Fc fragment of infliximab plays a pivotal role in impeding T cell activation, thus limiting the efficacy of TNF blockade in combination with ICI.

## Discussion

TNFi have been widely used to treat irAEs, such as colitis, in cancer patients undergoing ICI treatment^2^. There is a controversy regarding the pharmacodynamic and clinical impact of TNFi in combination with ICI in cancer patients^26-29^. Although infliximab effectively treats ICI-induced colitis, suggesting an anti-inflammatory impact, three different studies have found that it does not hinder the response to ICI treatment^26-28^. However, a retrospective analysis of a large cohort of ICI-treated advanced melanoma patients suggests that managing AEs with a combination of infliximab and corticosteroids is associated with shorter overall survival (OS) than in patients treated with corticosteroids alone^29^.

Herein, we present evidence that the co-administration of a tritherapy comprising TNFi (certolizumab or infliximab) with ipilimumab and nivolumab is safe for patients with advanced melanoma. However, depending on the TNFi used, clinical outcomes differed, with patients in the infliximab and certolizumab cohorts demonstrating better tolerability and anti-tumor activity, respectively. This is consistent with our observations during the safety assessment part of the TICIMEL clinical trial^11^. Overall, both tritherapies increased the number of circulating CD4^+^ and CD8^+^ T cells, promoting their maturation and proliferation, as well as increasing the proportion of Th1-like cells, as observed in patients treated with nivolumab and ipilimumab alone^12^. Both tritherapies also promoted an increase in proportions of circulating CD8 T cells targeting MelanA, a melanocyte-derived antigen that can be expressed by melanomas. This observation is important because it shows that TNF blockade does not prevent the potentiation of tumor-specific responses in patients. CITE-Seq analyses of pre- and on-treatment CD8^+^ and CD4^+^ T cells show that ipilimumab and nivolumab increase the expression of molecular signatures associated with T cell activation and IFN signaling in effector cells, particularly HLA-DR^+^ CD8^+^ T cells. Interestingly, co-administration of certolizumab amplifies this phenomenon, whereas infliximab reduces it. The proportion of circulating HLA-DR^+^ CD8^+^ T cells has previously been shown to increase in the blood of cancer patients (including those with melanoma or non-small cell lung carcinoma) treated with ICI^30,31^. This phenomenon has even been linked to a better response to ICI in renal cell carcinoma^32^. In addition, we observed an increase in the proportion of CD8 T cells producing IFN-γ in patients treated with either bitherapy or the certolizumab tritherapy. This increase was associated with an objective clinical response in the certolizumab tritherapy cohort. However, no such increase was observed in patients receiving tritherapy with infliximab, further indicating that the two TNFi have distinct pharmacodynamic impacts on CD8 T cells. Thus, it is possible that infliximab hinders the expansion of clinically active CD8 T cells. In patients affected with autoimmune diseases, infliximab triggered a transient reduction in circulating CD8 T cell subsets (i.e., CD8^+^ TEMRA) that expressed membrane-bound TNF and exhibited antimicrobial activity against *M. tuberculosis*^*33*^. Although we observed an increase in the proportion of CD8^+^ TEMRA cells in the infliximab cohort at W6 following treatment induction, we cannot rule out the possibility that infliximab transiently depletes clinically active CD8 T cells. This could explain why patients co-administered with infliximab and ICI tolerated treatment better but showed less efficacy than those in the certolizumab cohort.

Circulating Tregs increased in the blood of patients regardless of the therapy received. However, we observed an increase in the expression of genes related to IFN-γ signaling in Tregs from patients treated with the bitherapy, but not in those receiving any of the tritherapies. Thus, TNF signaling may contribute to the conversion of Tregs into STAT1-expressing Tregs, alongside IFN-γ produced by effector T cells^34^. Another hypothesis is that TNF, possibly in conjunction with TGF-β, plays a role in converting Th1 cells into Tregs. These Tregs have been shown to potently inhibit Th1 cell responses^34^, impairing anti-cancer immunity and the response to anti-PD-1 in the B16-OVA mouse melanoma model^35^. Since a subset of potent immunosuppressive Tregs express TNFR2^14-16^, it is possible that TNFR2-dependent TNF signaling contributes to the survival and biological activity of Tregs in patients with advanced melanoma. Our observations in preclinical mouse melanoma models reinforce this hypothesis, demonstrating that the combination of anti-TNF with ICI further reduces the proportion of Tregs infiltrating tumors, in addition to the impact of ICI on decreasing Tregs in tumors, possibly as a consequence of Treg depletion by anti-CTLA-4^36^. In wild-type mice, the reduced infiltration of Tregs in tumors, alongside the reduced expression of various immunosuppressive molecules on TILs (e.g. TIGIT, CD39, TIM-3 and PD-L1) is consistent with studies showing the key role played by TNF signaling in the expression of immunosuppressive molecules such as TIM-3 and PD-L1^9,37^. Interestingly, the proportion of exhausted T cells that co-express various immunosuppressive molecules, which are likely to contribute to resistance mechanisms to ICI, was significantly reduced upon tritherapy. This suggests that TNF signaling is involved in the exhaustion of CD8^+^ TILs. In addition, we observed less cell death in CD8^+^ TILs, which is consistent with the hypothesis that TNF signaling contributes to the AICD in CD8^+^ T cells^8-10,38,39^. To gain further insight into the impact of TNF blockade using certolizumab or infliximab in combination with ICI, we conducted experiments on preclinical mouse melanoma models in TNF and TNFR2-humanized mice. Certolizumab enhanced the efficacy of ICI, whereas infliximab did not. As observed in wild-type mice with the rat IgG2a anti-TNF, TNF blockade with certolizumab, but not with infliximab, reduced Treg infiltration into tumors and the exhaustion of CD8^+^ TILs. In mice treated with the tritherapy containing certolizumab, we observed a reduced expression of two key transcription factors: FOXP3, which is involved in the development and biological function of Tregs^40^, and TOX, which is involved in the exhaustion of CD8 T cells^41,42^. Full IgG1 anti-TNF (e.g. adalimumab) has been shown to trigger paradoxical membrane TNF-TNFR2 stimulation, leading to the expansion of Tregs in rheumatoid arthritis^42^. Herein, the proportion of dead cells among tumor-infiltrating Tregs decreased significantly in mice treated with the tritherapy containing infliximab compared to mice treated with the tritherapy containing certolizumab.

We hypothesized that the difference between the two TNFi is due to the fact that infliximab has a human IgG1 Fc fragment. This may make it more immunogenic in mice than certolizumab and affect immune responses. To reduce infliximab’s immunogenicity, we murinized it and demonstrated that opposite results can be obtained depending on the type of Fc fragment. Administering mouse IgG1 Fc infliximab, which is considered as functionally equivalent to human IgG4 Fc infliximab^24^, enhanced the efficacy of ICI, alongside a reduction in tumor-infiltrating Tregs and exhaustion of CD8^+^ TILs, as was observed with certolizumab. In contrast, administering mouse IgG2a Fc infliximab, which is functionally equivalent to human IgG1 infliximab^25^, did not enhance the efficacy of ICI. Moreover, we observed greater cell death in CD8^+^ TILs in mice treated with the tritherapy containing mouse IgG2a Fc infliximab or human IgG1 Fc infliximab than in those treated with the tritherapy containing mouse IgG1 Fc infliximab. As TNFR1 and TNFR2 signaling has been shown to trigger AICD in CD8^+^ T cells^8-10,38,39^, mouse IgG2a Fc infliximab or human IgG1 Fc infliximab could drive paradoxical cell death in activated effector T cells.

Given that infliximab (i) binds to the activating receptors FcγRI, FcγRIIa, and FcγRIIIa, and the inhibitory receptor FcγRIIb when complexed with TNF^44^, (ii) induces antibody-dependent cell cytotoxicity (ADCC)^44^, and (iii) binds to C1q, inducing complement-dependent cytotoxicity (CDC) in cells expressing membrane TNF^44^, these biological properties can also explain the differences with certolizumab, which lacks an Fc fragment. A systematic comparison of the binding properties of IgG isotypes to mouse FcγR revealed that mouse IgG2a and human IgG1 have a similar ability to interact with the four mouse FcγR (e.g. I, IIB, III and IV) when complexed with their antigens. However, mouse IgG1 only binds to mouse FcγRIIB and mouse FcγRIII^45^. Interestingly, rat IgG2a does not bind to mouse FcγRs, except for a weak interaction with mouse FcγRIII^45^. This may explain why the efficacy of ICI therapy is enhanced by rat IgG2a anti-TNF in wild-type mice and by mouse IgG1 infliximab in TNF and TNFR2-humanized mice, but not by mouse IgG2a infliximab or human IgG1 infliximab.

Collectively, our study demonstrates that TNF contributes to the resistance mechanisms and irAEs in advanced melanoma patients undergoing ICI therapy. Importantly, not all TNFi have the same pharmacodynamic and clinical impact, so they must be carefully selected based on clinical expectations. Therefore, infliximab and, probably the other TNFi with an IgG1 Fc fragment, are better suited for treating irAEs. In contrast, TNFi without an IgG1 Fc fragment, such as certolizumab, are preferable for improving the effectiveness of ICI therapy.

## Methods

### TICIMEL study design and participants

TICIMEL is an open-label phase 1b clinical trial assessing as a first objective the safety and tolerability of combining a TNF blocker (certolizumab or infliximab) with nivolumab and ipilimumab to treat patients with non-resectable locally advanced and/or metastatic melanoma. The secondary objective of TICIMEL was to assess treatment efficacy. This trial, which was conducted at the Oncopole Claudius Regaud (OCR) (France) was composed of a safety phase [2018-2019]^11^ during which 6 and 8 patients were treated with ipilimumab, nivolumab and infliximab or ipilimumab, respectively. During the course of the expansion phase, 12 and 6 additional patients have been enrolled in the certolizumab and infliximab cohorts, respectively. We report here the final results of the study.

Criteria for patients’ recruitment followed the previously published guidelines^11^. Briefly, eligible patients had histologically-proven Stage IIIc-IV unresectable locally advanced and/or metastatic melanoma with documented BRAFV600 status; were aged > 18 years and had an Eastern Oncology Group Performance status (ECOG) of 0 or 1, except one patient who had an ECOG score of 2. Key exclusion criteria were uveal melanoma and active intracranial disease. Before study entry, all patients gave their written, informed consent for participation in the clinical trial. This study was approved by the French committee for the protection of persons and the French drug agency [Agence Nationale de Sécurité du Médicament; date of approval

August 4, 2017 EUDRACT 2016-005139-34] and was registered under ClinicalTrials.gov, number NCT03293784. All study procedures were carried out in accordance with the International Council for Harmonization tripartite guideline on good clinical practice (Helsinki declaration). An Independent Data Monitoring committee (IDMC) monitored and evaluated data from the study.

### Patients and treatments (TICIMEL)

This trial was not randomized and patients were alternatively assigned to either one of both tritherapy cohort: ipilimumab + nivolumab + certolizumab (certolizumab cohort) or ipilimumab + nivolumab + infliximab (infliximab cohort). All patients were naïve of previous line of treatment (including adjuvant therapy with anti-PD-1 or with BRAFV600+MEK inhibitor; adjuvant therapy with interferon was allowed), excluding two patients from the certolizumab cohort and one patient from the infliximab cohort who were previously treated with adjuvant immunotherapy. Patients’ treatment regimen was designed as follow: 1 mg/kg of nivolumab every 3 weeks and 3 mg/kg of ipilimumab every 3 weeks for 4 doses, followed by 3 mg/kg of nivolumab every 2 weeks for cycle 5 and beyond. Patients enrolled in the certolizumab cohort, were treated with 400 mg sub-cutaneous certolizumab every 3 weeks for 3 doses; then 200 mg every 2 weeks for cycle 4 and beyond. Patients enrolled in the infliximab cohort were treated with 5 mg/kg of infliximab every 3 weeks for 3 doses, followed by 5 mg/kg of infliximab every 8 weeks. Tumor assessment was performed at baseline and every 12 weeks for 2 years or until disease progression by following the Response Evaluation Criteria in Solid Tumors (RECIST 1.1) using CT scan or MRI (17). Adverse events were graded according to the National Cancer Institute’s Common Terminology Criteria for AE (NCI CTCAE) version V4.03 and assigned as related or unrelated to study treatment by the investigator. Complete methods are available in the study protocol.

### Additional cohort and specimen collection

The bitherapy cohort includes twenty-five patients with histologically-proven Stage IIIc/IV metastatic and/or unresectable cutaneous melanoma patients (mucosal melanoma included) from the MELANFα clinical trial (NCT03348891)^12^. The treatment schedule of MELANFα patients included in this study was as follow: 1 mg/kg of nivolumab and 3 mg/kg of ipilimumab every 3 weeks for 4 infusions and then 240 mg or 480 mg of nivolumab every 2 or 4 weeks, respectively.

During the course of both the TICIMEL and MELANFα clinical trials, blood samples were collected at baseline (week 0, W0) and 6 weeks after the initiation of treatment (week 6, W6). Analyses were performed on fresh blood (EDTA) and/or isolated PBMC (Ficoll-Paque) and appropriately conserved in liquid nitrogen. The translational study comparing the immune responses in patients enrolled in TICIMEL and MELANFα has been registered on ClinicalTrials.gov as NCT05867004.

### Flow cytometry analyses

Fresh blood was incubated with antibody cocktails directed against lineage, maturation and differentiation markers of T cells before red blood cell lysis (BD Pharm Lyse™ 10X buffer [BD biosciences]), staining with a viability dye (Zombie NIR™, Biolegend) and fixation in 2% formal saline solution. Treg staining was performed on isolated CD4 T cells from patients’ PBMC (CD4 MicroBeads, human, Miltenyi Biotech), while T cell proliferation was assessed on total PBMC. Cells were first incubated with a cocktail of fluorescently labelled antibodies directed against membrane markers of each subpopulation and a viability dye followed by fixation and permeabilization using the “FOXP3 fixation/permeabilization buffer” (eBioscience, Thermofisher). Cells were then incubated with antibodies targeting intracellular markers before acquisition. The capacity of CD8 T cells to produce IFN-γ was assessed on CD8 T cells isolated from PBMC (CD8 MicroBeads, human, Miltenyi Biotech). All antibodies used in these analyses are listed below. CD8 T cells were polyclonally restimulated for 1h with a cocktail of Phorbol 12-myristate 13-acetate (PMA) and ionomycin (Cell stimulation cocktail, Thermofisher) before addition of a cocktail of Brefeldin A and monensin (Thermofisher) and incubation for 3 additional hours. To monitor the presence of MelanA-specific T cells in patients’ blood, CD8 T cells were isolated from pre-treatment (W0) and on-treatment (W6) PBMC using positive magnetic selection (MicroBeads, Miltenyi Biotec). The cells were stained with HLA-A2*0201 Melan-A (ELAGIGILTV) dextramers WB2162-PE (Immudex) at room temperature for 10 min. Then, the following fluorochrome-conjugated monoclonal antibodies were added: viability dye (Fixable Viability Dye eFluor™ 506; eBioscience, Thermo Fisher Scientific), anti-CD3 (UCHT1, AF700, BD Biosciences) and anti-CD8 (IT2.2 BV785, BioLegend). The cells were incubated for an additional 20 min at 4°C, washed, and fixed in 1% paraformaldehyde (Fisher Scientific) in PBS. Samples were acquired on a BD LSRFortessa X-20 (BD Biosciences), and data were analyzed using BD FACSDiva software. For each patient, PBMC analyses of both time points were performed in parallel.

| Target | Clone | Dye | Company |
| --- | --- | --- | --- |
| CD3 | UCHT1 | BV421 | Biolegend |
| CD3 | UCHT1 | APC | Biolegend |
| CD3 | UCHT1 | PerCP | Biolegend |
| CD4 | SK3 | BUV496 | BD Bioscience |
| CD8 | RPA-T8 | BUV395 | BD Bioscience |
| CD45RA | HI-100 | AlexaFluor 488 | Biolegend |
| CCR7 | G043H7 | PE-Cy7 | Biolegend |
| CXCR3 | G025H7 | BV711 | Biolegend |
| CCR6 | G034E3 | BV605 | Biolegend |
| CCR4 | L291H4 | BV421 | Biolegend |
| CD25 | BC96 | PECy7 | Biolegend |
| FOXP3 | PCH101 | AlexaFluor 700 | ThermoFisher Scientific |
| IFN-γ | 4S.B3 | PECy7 | Biolegend |
| Ki67 | B56 | AlexaFluor 647 | BD Pharmingen |
| Zombie NIR | na | APC-Cy7 | Biolegend |

### PBMC preparation for CITE-Seq

PBMC, isolated with Ficoll-Paque from W0 and W6 fresh blood were prepared within two hours following patients’ blood sampling and appropriately frozen in liquid nitrogen until further use. PBMC were incubated for 10 minutes at 4°C with Human Trustain FcX Fc blocking reagent (Biolegend) using the Cell Staining Buffer (Biolegend). Barcoded TotalSeq-A antibodies (Biolegend) (see table below) diluted in Cell Staining Buffer have been added for 30 minutes at 4°C, before cell washing, filtering (40um strainer, Falcon) and resuspension in 1X PBS before CITE sequencing. CITE-Seq was performed by dedicated engineers of the technological core facility of the Cancer Research Center of Toulouse. All samples of a given patient were handled and sequenced at the same time.

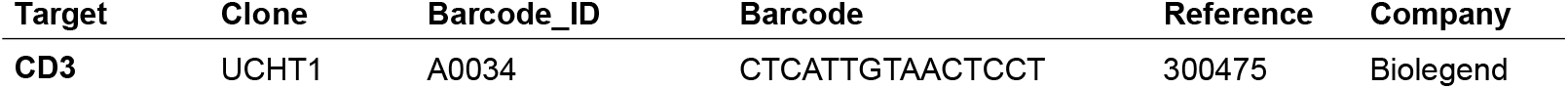
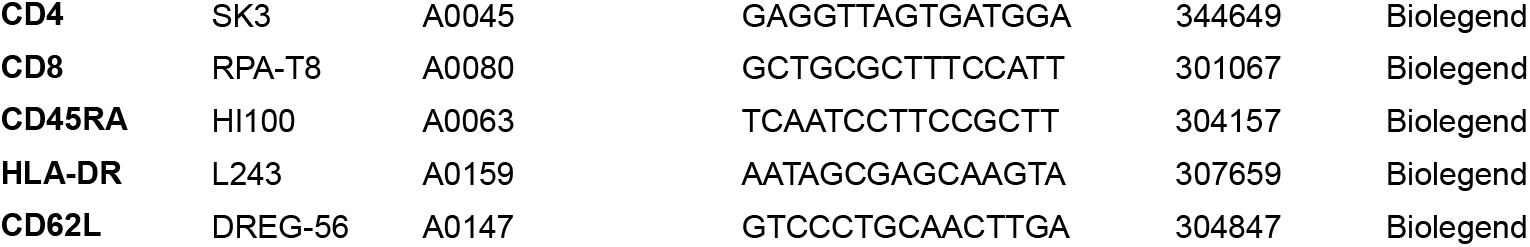

### CITE-Seq analyses

Analyses were performed as previously described^12^. Briefly, single-cell libraries (3’ gene expression and antibody-derived tags) were generated using Chromium Instruments and Chromium Single Cell 3⍰ Library & Gel Bead Kit v3 according to the manufacturer’s protocol (10× Genomics and Biolegend). For each sample, 15,000 cells were injected and library size and quality were confirmed on Fragment Analyzer (Agilent Technologies). The libraries were sequenced on a NextSeq 550 (Illumina) or a NovaSeq 6000 (Illumina) in paired end sequencing 28 bp (read1) × 91 bp (read2) and 8bp single index.

### Bioinformatic analyses

#### CITE-Seq on patients’ PBMC

##### Pre-Processing

CITE-Seq data were aligned to the human transcriptome (GRCh38-2020-A) using Cell Ranger version 7.1 (Include introns⍰=⍰T). The single-cell transcriptomics data from the TICIMEL and MELANFα clinical trials were processed using Seurat (v.5.0.0). For each sample, low quality cells were removed by assessing the number of detected genes versus the proportion of mitochondrial RNA for each sample (minimal number of genes per cell: 200; proportion of mitochondrial RNA: 6-12,5%). The samples were then merged bringing the total of cells to 348,427 and integrated with the Canonical Correlation Analysis (CCA) method. The clustering was based on Seurat functions and was performed using Unified Manifold Projection and Approximation (UMAP). We also relied on the Clustree (v.0.5.1) package to select the optimal cluster resolution based on stability. To help identify the immune cell clusters, the protein layer (ADT) was also added to the Seurat object. We identified 9 clusters of immune cells including cluster of CD4 and CD8 T cells, which were further subsetted and analyzed in depth.

##### Selection of CD4 and CD8 T cells

Different populations were identified and investigated using the Single Cell Signature Explorer tool (SCSE)^46^, which allows interactive scRNAseq UMAP data exploration. Scores were computed based on signatures including relevant gene markers such as *CD3* (*CD3D*; *CD3G*) and *CD8* (*CD8A*; *CD8B*) for the first selection of cells and previously published signatures for the functional enrichments^47^. We used the Merger tool to map the cells on the UMAP and then used the Virtual Cytometer to determine relevant thresholds for cell subtype definition. The selection of CD4 and CD8 cells required 4 different signatures for exclusion of non-T cells and validation of T cell identity:

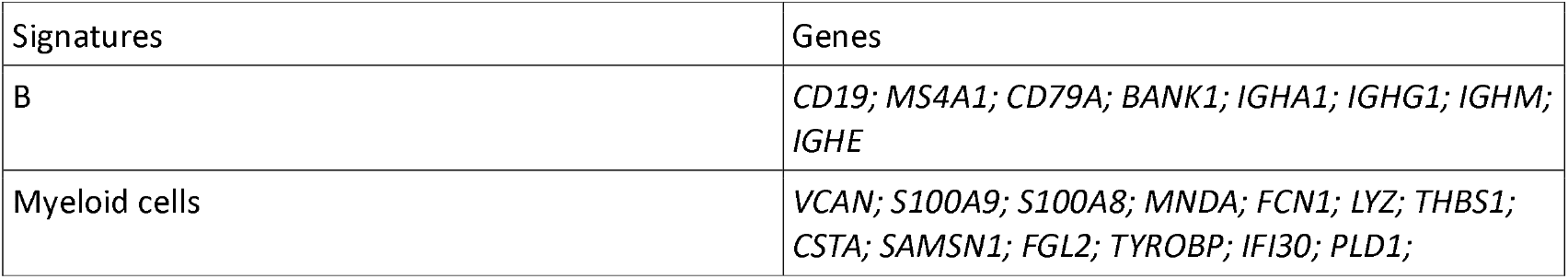
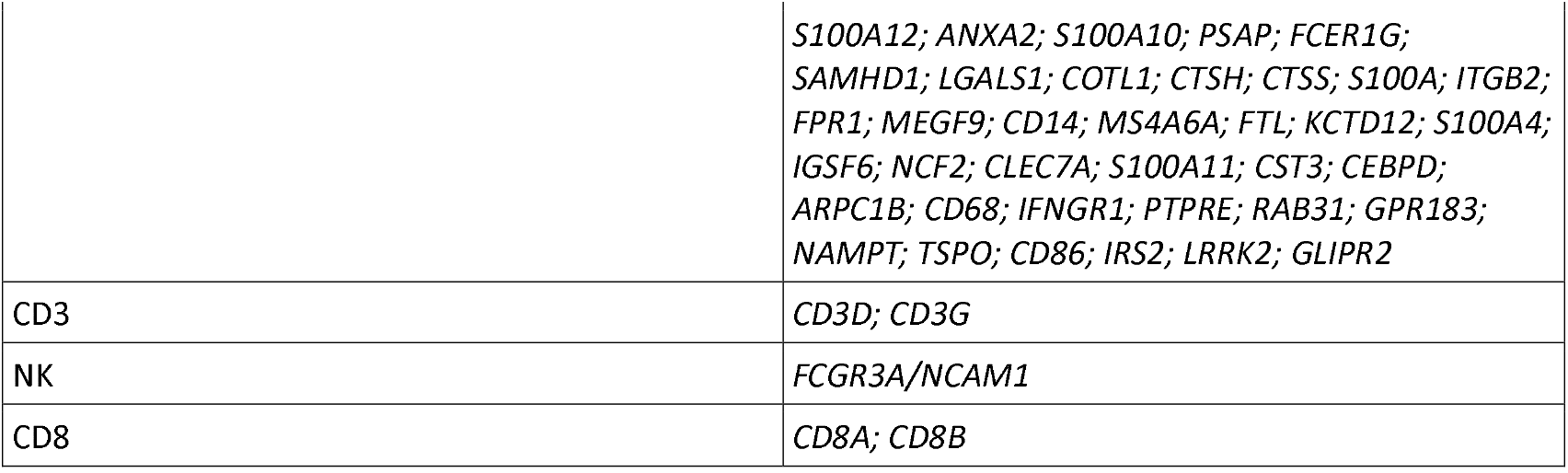

To complete the selection of CD8 cells, those expressing the *CD4* gene layer were excluded along with remaining contaminating cells expressing: *NCAM1, NCR1, IGHM, IGHG1, IGHA1*. For CD4 T cells and Tregs, we also further excluded cells expressing *FCGR3A, NCR1, CD8A* and *CD8B*. Tregs were selected from the CD4 T cells through the use of SCSE tools with thresholds applied on cells expressing *FOXP3* and *IL2RA* genes.

##### Analysis

The SCP (v.0.5.6) package including slingshot was used to perform the trajectory analysis on the CD8 cells after exclusion of cluster 5 (MAIT cells). Trajectories were calculated using the slingshot function, with cluster 3 (Naive cells) as a starting point.

Differential gene expression analyses were performed using the Seurat package. Pathway enrichment analyses were performed using the Over Representation Analysis (ORA) method with the “fora” function of the fgsea package (v.1.18.0). All pathways were extracted from the Msig database. For the following analysis the 10 most enriched pathways for each treatment were selected, based on their adjusted p-value. The Fold Enrichment (FE) measure, as shown in Fig. 2f, reflects the over-representation of genes or pathways in a list of genes of interest and was calculated using the following formula:

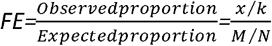

x: number of genes significantly differentially expressed enriched in the pathway, k: number of all of the significantly differentially expressed genes, M: size of the gene set and N: size of the universe.

Heatmaps in Fig. 2g and Extended data Fig. 4b show genes linked to pathways identified in Fig. 2f and Extended data Fig. 4c, respectively. Depicted genes were filtered so that the log2 fold change (W6/W0) of the expression of any given gene, calculated per patient, had to be significantly different between at least 2 treatment cohorts. Inference of transcription factor activity was performed using the DecoupleR (v.2.8.0) pipeline on pseudo-bulk profiles generated by summing the normalized expression values per cluster, treatment, response and timepoint. Wilcoxon tests with no correction were used for statistical analyses. For a given cluster, only patients with at least 10 cells at both time points are represented in the heatmaps.

#### Analysis of bulk RNA-Seq on mouse tumors

Raw gene count matrices were used as input for differential expression analysis. Genes with fewer than 10 reads across all samples were filtered out to remove low-abundance transcripts. Differential gene expression analysis and normalization were performed using the DESeq2 package (version 1.48.2) in R (version 4.5.1). Gene Ontology (GO) enrichment analysis of Biological Processes (BP) was then carried out with the gseGO() function from the clusterProfiler package (version 4.16.0), using genes ranked by log2 fold change. Pathways with an adjusted p-value < 0.01 were visualized using the aPEAR package (version 1.1.1). Similarity between pathways was evaluated using the Jaccard index (default) and used to detect clusters of redundant pathways through the Markov clustering algorithm (default). Network analysis was then applied to identify the pathway best representing each cluster using the default PageRank algorithm (default). Clusters of interest (e.g., “positive regulation of leukocyte/lymphocyte activation”) were further summarized using the rrvgo package (version 1.20.0) to reduce redundancy and facilitate biological interpretation.

### Mice

Wild-type (WT) C57BL/6 mice were purchased from Janvier Labs. hTNFKIxhTNFR2KI mice have been previously described^17^ and were kindly provided by Dr. Ilgiz Mufazalov (Mainz, Germany). For each experimental condition, six-to ten-week-old female mice were used. All mice were maintained in our animal facility under SOPF conditions with a maximum of five mice per cage (US006 CREFRE-Inserm/UPS), which is accredited by the French Ministry of Agriculture (accreditation number A31555010). All animal experiments were conducted in accordance with the French and European regulations for the care and protection of laboratory animals, and were approved by the local ethics committee and the Institutional Animal Care and Use Committee.

### Cell lines

Murine melanoma cell lines B16-OVA and B16-F10 were cultured in DMEM (Gibco, #61965-026) supplemented with 10% inactivated fetal bovine serum (FBS) (Pan Biotech, P30-3306) and 10% penicillin/streptomycin (Pan biotech, P06-07100), under standard culture conditions. The YUMM1.7 melanoma cell line (Yale University Mouse Melanoma) was cultured in Opti-MEM medium (Gibco, #31985-047) supplemented with 10% FBS and 10% penicillin/streptomycin. All cell lines were routinely tested for mycoplasma contamination and found to be negative.

### Tumor models

For tumor implantation, 3 × 10^5^ cells (for B16-OVA or YUMM1.7) or 1 × 10^5^ cells (for B16-F10), suspended in PBS, were injected intradermally. Mice were randomized into experimental groups once tumors became palpable. The tumor size was measured three times per week using a caliper, and the tumor volume was calculated as V (Volume) = 0.52 × length × width^2^. The generation of tumor growth curves was facilitated by GraphPad Prism, while R was employed to generate waterfall plots, thereby illustrating the log_2_-transformed percentage change in tumor volume between days 6 and 28. For mice that died before day 28, the final recorded tumor measurement was used for analysis.

### Animal treatment protocol

Wild-type C57BL/6 mice were treated with a combination of anti-PD-1 and anti-CTLA-4 antibodies, and a murine anti-TNF antibody when indicated. During the induction phase, the mice received an initial cycle of 200 µg each of the three antibodies, followed by three subsequent cycles of 200 µg of anti-PD-1 and 100 µg of anti-CTLA-4 per injection, with or without 200 µg of anti-TNF. During the maintenance phase, the mice received three cycles of 200 µg of anti-PD-1 alone or in combination with 200 µg of anti-TNF. Humanized hTNF × hTNFR2 knock-in (hTNF-KI × hTNFR2-KI) mice received 200 µg each of anti-PD-1, anti-CTLA-4, and the indicated anti-TNF antibody (certolizumab, infliximab, or modified infliximab constructs) for four induction cycles. During the maintenance phase, the mice received three cycles of 200 µg each of anti-PD-1 and the corresponding TNFi.

### Treatments

Anti-PD-1 (RMP1-14), anti-CTLA-4 (9H10), anti-TNF (XT3.11) and modified infliximab (infliximab mouse IgG1 or mouse IgG2a) were purchased from BioXcell. Certolizumab (Cimzia 9342601, UCB) and infliximab (Inflectra 9398352, Pfizer) were purchased from the IUCT-O pharmacy.

### Flow cytometry analyses in mice

Tumors and tumor-draining lymph nodes were collected at the indicated timepoints. Lymph nodes were mechanically dissociated, while tumors were processed using mechanical and enzymatic digestion using the Tumor Dissociation kit (Miltenyi) to obtain single-cell suspensions. CD45^+^ cells from tumor cell suspensions were isolated using the CD45 (TIL) MicroBeads, Mouse (Miltenyi). Cells were then incubated with the Mouse TruStain FcX and True-Stain Monocyte Blocker (Biolegend) before staining with antibodies targeting membrane proteins, fixation/permeabilization using the “FOXP3 fixation/permeabilization buffer” (eBioscience, Thermofisher) and intracellular staining to characterize T cell subsets within tumors and lymph nodes. The following antibodies and reagents were used: CXCR3 (Clone CXCR3-173 BD Biosciences); LAG-3 (Clone C9B7W BD Biosciences); PD-1 (Clone 29F.1A12 BioLegend); TIM-3 (Clone RMT3-23 BioLegend); TIGIT (Clone 1G9 BioLegend); KLRG1 (Clone 2F1/KLRG1 BioLegend); CD103 (Clone 2E7 invitrogen); CD3 (Clone 17A2 Invitrogen); CD4 (Clone GK1.5 BioLegend); PD-L1 (Clone MIH5 Invitrogen); CD69 (Clone H1.2F3 BioLegend); CD44 (Clone IM7 BioLegend); CD45.2 (Clone 104 BioLegend); CD39 (Clone Y23-1185 BD Biosciences); CD27 (Clone LG.3A10 BD Biosciences); CD8 (Clone 53-6.7 BD Biosciences); CD25 (Clone PC61 BioLegend); CD62L (Clone MEL-14 BioLegend); SLAMF6/Ly108 (Clone 13G3-19D Invitrogen); MHC Class II (Clone M5/114.15.2 BioLegend); CD127 (Clone A7R34 Invitrogen); BTLA (Clone 6A6 BioLegend); CD226 (Clone 10 E 5 BioLegend); Granzyme B (Clone N4TL33 Invitrogen); BCL-6 (Clone K112-91 BD Biosciences); FoxP3 (Clone FJK-16s invitrogen); Tbet (Clone 4B10 BD Biosciences); Tox (Clone NAN448B BD Biosciences); Ki67 (Clone B56 BD Biosciences); CTLA-4 (Clone UC10-4B9 BioLegend); CD45 (Clone 30-F11 BD Biosciences); Zombie NIR (#423105 Biolegend). Acquisition was performed on the Aurora 5L spectral cytometer (Cytek). For cytokine staining, cells were restimulated for 4h with cell stimulation cocktail plus protein transport inhibitors (#00-4975-03 ThermoFischer). The following antibodies and reagents were used: LIVE/DEAD Fixable Aqua cell satin kit (Thermo Fisher, L34965); CD3 (Clone 17A2 BioLegend); CD4 (Clone GK1.5 BD Biosciences); CD8 (Clone 53-6.7 BioLegend); T-bet (Clone 4B10 Invitrogen); TNF (Clone MP6-XT22 BD Biosciences); GzmB (Clone GB11 BioLegend); IFN-γ (Clone XMG1.2 BD Biosciences). For the analysis of T cells infiltrating B16F10 melanomas, tumors were processed as described above and total cell suspensions were first incubated with Mouse TruStain FcX (Biolegend), then the following antibodies targeting membrane proteins: CD45 (Clone 30-F11 BD Biosciences), TCRβ (Clone H57-597 BD Biosciences); CD4 (Clone GK1.5 BD Biosciences); CD8 (Clone 53-6.7 BioLegend), LIVE/DEAD Fixable Aqua cell satin kit (Thermo Fisher, L34965). Cells were then fixed and permeabilised as described above and stained with antibodies targeting Foxp3 (Clone FJK-16s invitrogen). Data acquisition was performed on a BD Fortessa X-20, and data of both sets of experiments was analyzed using OMIQ and FlowJo software.

### Bulk RNA-Seq

Tumors were collected at the indicated timepoints and mechanically dissociated using Lysing Matrix D tubes (MP Biomedicals) using the Precellys Evolution homogenizer (Bertin Technologies). Total RNA was extracted using the RNeasy Midi Kit (QIAGEN) according to the manufacturer’s instructions. RNA integrity and concentration were assessed prior to library preparation (Fragment Analyzer, Agilent). Bulk RNA sequencing was performed by the technological core facility of the Cancer Research Center of Toulouse (CRCT) using Illumina Stranded mRNA Prep Ligation kit. The libraries were sequenced on a NextSeq 550 (Illumina) or a NovaSeq 6000 (Illumina, 2×100bp).

### Statistical analysis

#### Clinical data from TICIMEL and MELANFα trials

Descriptive statistics were used to summarize demographics, tumor characteristics and treatments. Categorical variables were summarized using frequencies and percentages and continuous variables using median and range (minimum-maximum). AEs were described using frequencies and percentages according to toxicity grade for each event. Response data were described using frequencies and percentages and tumor changes were summarized graphically. Progression-free survival was calculated from study inclusion to progressive disease or death, whichever occurs first, and estimated using the Kaplan-Meier method for each cohort. Paired continuous data between W0 and W6 were compared using the Wilcoxon matched-pairs signed-rank test. All statistical tests were two sided and p-values <0.05 were considered significant. All statistical analyses were performed using STATA software version 16.

#### Murine data

Statistical analyses were performed using GraphPad Prism. Data were first assessed for normality, and appropriate statistical tests were applied accordingly. For normal distributed data, comparisons between two groups were performed using an unpaired t-test, and comparisons among multiple groups were performed using a one-way ANOVA with appropriate post hoc tests. For non-normally distributed data, the corresponding non-parametric tests were used. Data are presented as mean ± s.e.m. Differences were considered significant at P < 0.05 (*), P < 0.01 (**), P < 0.001 (***) or P < 0.0001 (****).

## Acknowledgments

We thank the patients who participated in this study; the clinical research staff, including all sub-investigators, nurses, clinical research associates, data managers and secretaries. We thank Bristol-Myers Squibb (BMS) for funding the clinical trial and part of the ancillary study. We thank Fondation ARC pour la recherche sur le cancer, Institut National du Cancer (INCa), Labex Toucan, Cancéropôle Grand Sud-Ouest (GSO) and Institut Universitaire du cancer Toulouse-Oncopole (IUCT-O), and Ligue Régionale contre le Cancer Midi-Pyrénées for funding part of the ancillary study. This project has received funding from the European Union’s Horizon 2020 Research and Innovation Programme under grant agreement No. 964264 (TRANSCAN-3). We thank the Fondation Toulouse Cancer Santé (FTCS), the Fondation pour la Recherche Médicale (FRM) and Région Occitanie (Pyrénées-Méditéranée) for covering the salary supports of MV, SB and BJ. This study was partially supported by the grant EUR CARe N°ANR-18-EURE-0003 in the framework of the Programme des Investissements d’Avenir. We thank the CARe Graduate School for the support of Tania Margarido Pereira, Mathieu Virazels, Benjamin Jung and Matthieu Genais. VP acknowledges the Chair of Bioinformatics in Oncology of the CRCT (INSERM; Fondation Toulouse Cancer Santé and Pierre Fabre Research Institute). We thank the technology cluster of the CRCT, in particular Manon Farcé, Emeline Sarot and Carine Valle for their help with flow cytometry analyses and RNA sequencing. We thank the members of the Oncopole Claudius Regaud (OCR) who contributed to the design and management of the study.

## Protocol

The full protocol for this trial is available.

## Ethical considerations

All study procedures were performed in accordance with the International Council for Harmonization tripartite guideline for good clinical practice (Declaration of Helsinki). All patients enrolled in TICIMEL (NCT03293784) provided their written informed consent prior to study entry.

## Authors’ contributions

TMP, MV, BJ, SB, LB, LL, CC and AM performed the experiments and analyzed the data. CC, AM and BS jointly supervised the project and wrote the paper. All the authors edited the paper. TMP, MV and BJ contributed to the thinking behind certain parts of the project and the experimental choices, contributed to the structure of the article, participated in writing various sections of the manuscript (methods, figures, captions), proofread and edited the manuscript, and reworked the discussion. AL performed the biostatistics and edited the paper. TF wrote the methodology of the clinical trial protocol and supervised the biostatistics. MV, BJ, TMP, SB, CMS and AM contributed to the experiments of the ancillary part. EL, MG and BJ performed the bioinformatics analyses, and JPC and VP supervised. CP, VS and NM contributed to patient enrollment and clinical follow-up. MA, NAA, LM and CC contributed to the design of the ancillary part and interpretation of the immunological data. JPD and MM contributed to the writing of the clinical protocol. SN provided mice humanized for TNF and TNFR2 and contributed to data interpretation. IM advised about protocols and contributed to data interpretation. BS and NM designed the clinical trial. BS and AM designed and supervised the ancillary part. NM was the principal investigator of the TICIMEL clinical trial.

## Additional information

Claudius Regaud Oncopole (OCR) and Cancer Research Center of Toulouse (CRCT) designed the TICIMEL clinical trial (NCT03293784). OCR and CRCT wrote the TICIMEL protocol, which was reviewed and approved by Bristol Myers-Squibb (BMS). The study was sponsored by Oncopole Claudius Regaud. The decision to submit the paper for publication was made jointly by OCR and CRCT. The corresponding author confirms that he had full access to all the data in the study and takes final responsibility for the decision to submit for publication.

## Data availability statement

The anonymized derived data underlying the results of this article will be made available, starting 12 months and ending 5 years after the publication of this article in a peer-reviewed journal, to any investigator who signs a data access agreement and submits a methodologically sound proposal. Single-cell RNA-Seq data and analysis scripts will be deposited on GEO. Additional information is available upon request to the corresponding author.

## Supplementary material

**Extended data Table 1:** Study Treatment.

|  | Total<br>(N = 32) | Certolizumab<br>(N = 20) | Infliximab<br>(N = 12) |
| --- | --- | --- | --- |
| **Reason for treatment discontinuation (n=32)** |  |  |  |
| Maximum duration (1 year) | 3 (9.4%) | 1 (5.0%) | 2 (16.7%) |
| Toxicity | 17 (53.1%) | 13 (65.0%) | 4 (33.3%) |
| Progression | 8 (25.0%) | 3 (15.0%) | 5 (41.7%) |
| Death | 2 (6.2%) | 1 (5.0%) | 1 (8.3%) |
| Investigator decision | 1 (3.1%) | 1 (5.0%) | 0 (0.0%) |
| Other reason | 1 (3.1%) | 1 (5.0%) | 0 (0.0%) |
| **Total Treatment Duration (Months) (n=32)** |  |  |  |
| Median | 1.41 | 0.79 | 2.10 |
| (Range) | (0.03;11.99) | (0.03;11.76) | (0.03;11.99) |
| **Number of injections for Nivolumab(IV) (n=32)** |  |  |  |
| Median | 3.0 | 2.0 | 4.0 |
| (Range) | (1.0;24.0) | (1.0;23.0) | (1.0;24.0) |
| **Number of injections for Ipilimumab(IV) (n=32)** |  |  |  |
| Median | 3.0 | 2.0 | 3.0 |
| (Range) | (1.0;4.0) | (1.0;4.0) | (1.0;4.0) |
| **Number of injections for Certolizumab(SC) (n=20)** |  |  |  |
| Median |  | 2.0 |  |
| (Range) |  | (1.0;14.0) |  |
| **Number of injections for Infliximab(IV) (n=12)** |  |  |  |
| Median |  |  | 3.0 |
| (Range) |  |  | (1.0;8.0) |

**Extended data Table 2:**
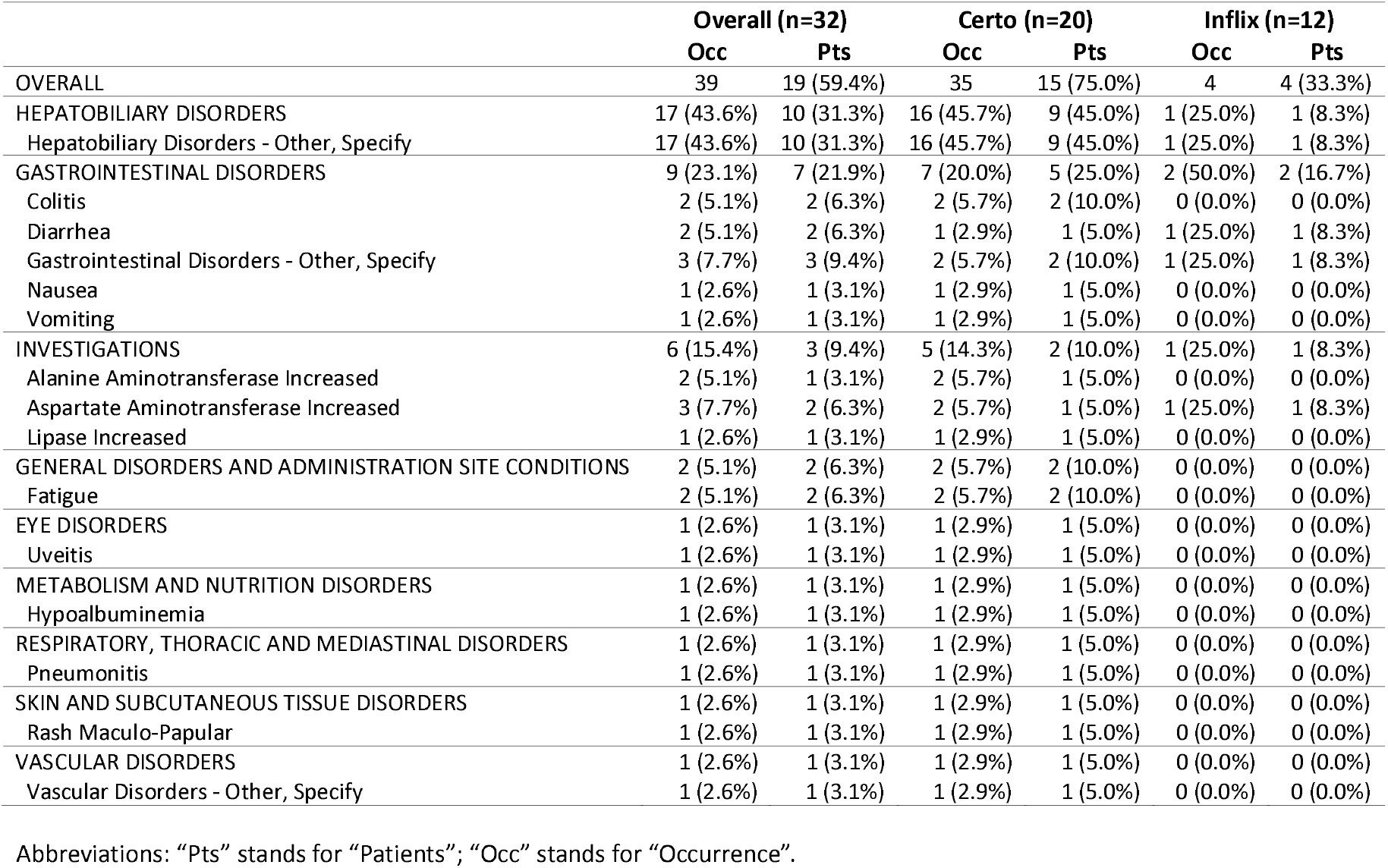
Incidence of Grade 3-5 related to study drug by SOC/PT.

## Legends to supplementary Figures

**Supplementary Figure 1. Description of the cohorts, patients’ clinical response, and circulating T helper subpopulations. a**, Table showing the analysis population of the TICIMEL trial. **b**, Swimmer plots showing the evolution of tumor burden as well as new event and treatment duration of patients treated with ipilimumab, nivolumab and certolizumab (left panel) or ipilimumab, nivolumab and infliximab (right panel). One patient in complete and one in partial response from the certolizumab and infliximab cohort, respectively, were not evaluable as per RECIST criteria at the first two tumor assessments. One patient from the certolizumab cohort and one from infliximab cohort progressed before the first tumor assessment. One patient from the certolizumab cohort and one patient from the infliximab cohort died before the first tumor assessment. x: patient in complete response despite 2 persistent lymph node lesions (<10 mm). **c**, Progression-free survival of patients treated with ipilimumab, nivolumab and certolizumab (blue) or ipilimumab, nivolumab and infliximab (red). d, Proportions of circulating Th2-like (CXCR3^-^CCR6^-^CCR4^+^); Th17-like (CXCR3^-^CCR6^+^) and Th17/1 (CXCR3^+^CCR6^+^) CD4^+^ cells between W0 and W6. Statistics: Wilcoxon matched-pairs signed-rank test.

**Supplementary Figure 2. Protein expression on circulating CD8 T cells and over-representation analysis. a**, Feature plots showing the expression of CD3, CD4, CD8, CD62L, CD45RA and HLA-DR proteins on CD8 analyzed by CITE-Seq. **b**, Proportion of cells in clusters 6 between W0 and W6 per cohort **c-e**, Differential gene expression analysis was performed comparing W6 and W0 cells from cluster 2 (separately on each treatment cohort), before over-representation analysis (ORA) on the lists of differentially expressed genes. The 10 most significantly deregulated pathways after ipi/nivo (b), ipi/nivo/certo (c) or ipi/nivo/inflix (d) in cells from cluster 2 are shown (n=8 patients per cohort).

**Supplementary Figure 3. Impact of ICI combined with certolizumab or infliximab on the molecular signature of effector CD4 T cells**. Peripheral blood mononuclear cells isolated from the blood of patients (8 patients per cohort) before treatment initiation (week 0, W0) and 6 weeks after the beginning of treatments (week 6, W6) was analyzed by CITE-Seq. **a**, Unified Manifold and Approximation and Projection (UMAP) of CD4^+^ T cells of all patients and time points, identifying 11 clusters (C0-C11) of CD4 T cells (9 clusters of CD4^+^ T cells and 1 cluster of platelets and 1 unidentified cluster). **b** and **c**, Dotplot showing the expression of key gene markers in the identified CD4 subclusters (proportion and average expression). **d**, Feature plots showing the expression of CD3, CD4, CD8, CD62L, CD45RA and HLA-DR proteins on CD4 analyzed by CITE-Seq. **e**, Proportion of cells from cluster 1 between W0 and W6 per treatment cohort. *p<0.05 Wilcoxon. **f**, A differential gene expression analysis was performed to compare W6 and W0 cells from cluster 1, separating each treatment cohort. Over-representation analysis (ORA) was then performed on the 3 lists of differentially expressed genes (DEG) to identify the 10 most significantly deregulated pathways after ipi/nivo, ipi/nivo/certo or ipi/nivo/inflix. The bar plot shows the fold enrichment of the 17 identified pathways in cells from cluster 1 after ipi/nivo; ipi/nivo/certo or ipi/nivo/inflix. **g**, Genes contributing to the identification of pathways depicted in “e” were identified. The genes, which were differentially modulated (log_2_ fold change (W6/W0) of the gene expression) between at least two treatment cohorts were selected and plotted on a heatmap. Gene induction or downregulation is shown per patient and treatment cohort. Left hand side: yellow squares identifying a significant difference in gene expression comparing two treatment cohorts at a time (p<0.05, Wilcoxon). **h**, Heatmap showing the modulation in the activity of IRF1, IRF2, IRF3, STAT1 and T-bet transcription factors (TF) in cluster 1 cells between W0 and W6 per patient and treatment cohort (DecouplR package). Left hand side: yellow squares identifying a significant difference between the evolution of TF activities comparing two treatment cohorts at a time (p<0.05, Wilcoxon; ns: non-significant).

**Supplementary Figure 4. Certolizumab and infliximab inhibit the expression of a gene signature related to IFN response in regulatory T cells upon ICI. a**, Representative staining (upper left) and proportions of regulatory CD4 T cells (CD25^+^FOXP3^+^) assessed on W0 and W6 PBMC from patients treated with ipi/nivo, ipi/nivo/certo or ipi/nivo/inflix. **b**, Regulatory T cells expressing both IL2RA and FOXP3 genes were subsetted from cluster 5 of CD4 T cells. Differential gene expression analyses were performed to compare W6 with W0 cells of each treatment cohort. All genes significantly modulated between W0 and W6 (log2 fold change [FC] (W6/W0) gene expression) in all three cohorts are shown on the heatmap. **c**, Boxplot showing the log2 fold change (W6/W0) of the expression of HDAC9, STAT1, PCCA and MIEN1 per treatment cohort. **d**, Over-representation analysis (ORA) was performed on the list of DEG obtained by comparing W6 and W0 gene signatures of circulating Treg from ipi/nivo-treated patients.

**Supplementary Figure 5. TNF blockade reinforces anti-tumor immunity and long-term protection in melanoma models. a**, Individual tumor growth curves of C57BL/6 wild-type mice injected intradermally with B16-OVA cells and treated with vehicle, anti–PD-1 plus anti– CTLA-4 (bitherapy), or the same combination plus anti-TNF (tritherapy). The number of complete responses is indicated above each group. **b**, Rechallenge experiment: mice that achieved complete responses (“tumor-free”) after tritherapy were reinjected with B16-OVA cells. Left, individual tumor growth curves following rechallenge compared with naïve control mice. Right, Kaplan–Meier survival analysis and the number of mice rejecting the secondary tumor challenge. **c**, Impact of the timing of anti-TNF administration on therapeutic efficacy. Mice were divided into four groups receiving either: bitherapy during the induction phase followed by anti–PD-1 alone (maintenance), bitherapy followed by anti–PD-1 plus anti-TNF, tritherapy followed by anti–PD-1 alone, or tritherapy followed by anti–PD-1 plus anti-TNF. Waterfall plots show the log_2_ percentage change in tumor volume between day 6 and day 28. **d**, Heatmap of selected immune-related genes (including Tbx21, Klrk1, Prf1, Ifng, and Gzmb) from bulk RNA-Seq of tumors from bi- and tritherapy–treated mice. **e**, Bubble plots depicting expression profiles within CD4^+^ TIL clusters identified in Figure 3, based on spectral cytometry. Circle size indicates the percentage of cells expressing a given marker, and color reflects normalized mean fluorescence intensity (z-score). **f**, Frequency of immunosuppressive FOXP3^+^CD39^+^TIGIT^+^ regulatory T cells among CD4^+^ TILs according to treatment. Representative gating strategies and plots for each treatment are shown on the left, with quantification on the right. **g**, Bubble plots showing marker expression profiles within CD8^+^ TILs clusters. **h**, Frequency of apoptotic (dead) CD8^+^ T cells according to treatment. Representative histograms of staining for each treatment are shown on the left, and quantification histograms are shown on the right. **i**, Expression of PD-1, TIM-3, and CD39 on CD8^+^ T cells. Representative gating strategies and plots for each treatment are shown on the left, with quantification histograms on the right. **j**, Distribution of CD226 expression levels (CD226^−^, CD226^mid^, and CD226^high^) among CD8^+^ TILs. Representative histograms for each treatment are shown on the left, and quantification histograms for CD226^−^, CD226^mid^, and CD226^high^ subsets are shown on the right. **k**, Bubble plots depicting marker expression in CD4^+^ T cell clusters from tumor-draining lymph nodes (TDLN). **l**, Bubble plots showing marker expression in CD8^+^ TDLN clusters. (n) Schematic representation of the therapeutic regimen in the B16-F10 melanoma model. C57BL/6 mice received vehicle, bitherapy, or tritherapy during the 4-cycle induction phase, followed by three maintenance cycles of anti-PD-1 alone or combined with anti-TNF starting on day 6. **m**, Schematic representation of the therapeutic regimen and analyses performed in the B16-F10 melanoma model. 1 x 10^5^ B16-F10 cells were injected intradermally and treated with 3 cycles of anti-PD-1 plus anti–CTLA-4 (bitherapy) alone or in combination with anti-mouse TNF. **n**, Individual tumor growth curves according to treatment group. **o**, Quantification of intratumor regulatory T cells at day 12 in each treatment group, shown as representative histograms and summary quantification. Data are presented as mean ± s.e.m. Survival curves were compared using the log-rank (Mantel–Cox) test. Comparisons between two groups were performed using unpaired two-tailed t-tests, and multiple comparisons were analyzed using one-way ANOVA followed by Tukey’s post hoc test. P values < 0.05 were considered significant (P < 0.05; *P < 0.01; **P < 0.001; ***P < 0.0001).

**Supplementary Figure 6. Certolizumab, but not infliximab, enhances the efficacy of anti– PD-1 + anti–CTLA-4 combination in TNF/TNFR2-humanized melanoma models.**

**a-c**, Experiments performed in hTNFKIxhTNFR2KI C57BL/6 mice bearing YUMM1.7 melanoma. **a**, Schematic representation of the therapeutic regimen. Mice were injected intradermally with 3 × 10^5^ YUMM1.7 cells and, starting on day 2 when tumors became palpable, received four induction cycles of anti–PD-1 plus anti–CTLA-4 (bitherapy) alone or in combination with certolizumab or infliximab (tritherapy), followed by three maintenance cycles of anti–PD-1 with or without the same anti-TNF antibody. **b**, Mean tumor growth over time according to treatment. **c**, Kaplan–Meier survival of mice according to treatment. d-o, Experiments performed in hTNFKIxhTNFR2KI mice bearing B16-OVA melanoma, as described in Figure 4. **d**, Bubble plots depicting marker expression profiles within CD4^+^ tumor-infiltrating lymphocytes (TILs) clusters identified in Figure 4, based on spectral cytometry. Circle size represents the percentage of cells expressing a given marker, and color indicates normalized mean fluorescence intensity (z-score). CD4^+^ TILs were classified into effector, regulatory and early activated subsets. **e**, Bubble plots characterizing CD8^+^ TIL clusters described in the UMAP analysis of Figure 4, corresponding to early activated, central memory, late activated and exhausted subsets. **f**, Left, representative gating strategy; right, quantification histograms showing the frequency of apoptotic (dead) FOXP3^+^ regulatory CD4 T cells according to treatment. **g**, Left, gating strategy; right, quantification histograms showing apoptotic conventional CD4^+^ T cells across treatment groups. h, Same as (g), but for CD8^+^ TILs. **i**, Left, UMAP projection of CD4^+^ T cells from tumor-draining lymph nodes (TDLN) identifying naïve, effector, and regulatory subsets, with treatment-stratified UMAPs. Right, histograms quantifying each CD4^+^ T cell subset per treatment. **j**, Bubble plots depicting marker expression within CD4^+^ TDLN clusters. **k**, Left, gating strategy; right, quantification histograms showing the frequency of Th1 cells (CD4^+^CD44^+^CD62L^−^CXCR3^+^T-bet^+^) among conventional CD4^+^ T cells. **l**, Left, UMAP projection of CD8^+^ TDLN clusters identifying naïve, effector, and regulatory subsets, with treatment-specific distributions; right, histograms showing subset frequencies. **m**, Bubble plots characterizing marker expression in CD8^+^ TDLN clusters. n, Left, gating strategy; right, histograms showing the frequency of proliferating PD-1^+^Ki67^+^CD8^+^ T cells across treatments. o,) Left, gating strategy; right, quantification histograms of Tpex-like CD8 T cells (TCF1^+^ SLAMF6^+^PD-1^+^TIM-3^−^) according to treatment. Data are presented as mean ± s.e.m. Survival curves were compared using the log-rank (Mantel–Cox) test. For tumor growth and flow cytometry analyses, comparisons between two groups were performed using unpaired two-tailed t-tests, and multiple comparisons were analyzed using one-way ANOVA followed by Tukey’s post hoc test or Kruskal–Wallis test where appropriate. P values < 0.05 were considered significant (P < 0.05; *P < 0.01; **P < 0.001; ***P < 0.0001).

**Supplementary Figure 7. Differential marker expression in CD4**^**+**^ **and CD8**^**+**^ **tumor-infiltrating lymphocyte subsets following TNF blockade with infliximab Fc variants**.

**a**, Bubble plots depicting marker expression profiles within CD4^+^ tumor-infiltrating lymphocyte (TILs) clusters identified in Figure 5, based on spectral cytometry. Circle size represents the percentage of cells expressing a given marker, and color indicates normalized mean fluorescence intensity (z-score). CD4^+^ TILs were classified into early activated, effector, and regulatory subsets. **b**, Bubble plots characterizing CD8^+^ TIL clusters described in the UMAP analysis of Figure 5, corresponding to short-lived effector, effector, exhausted and early activated subsets.

