## Supplementary figures and images for "TNF blockade with certolizumab improves the efficacy of anti-PD-1 and anti-CTLA-4 combination therapy for melanoma"

### Supplementary Figure 1

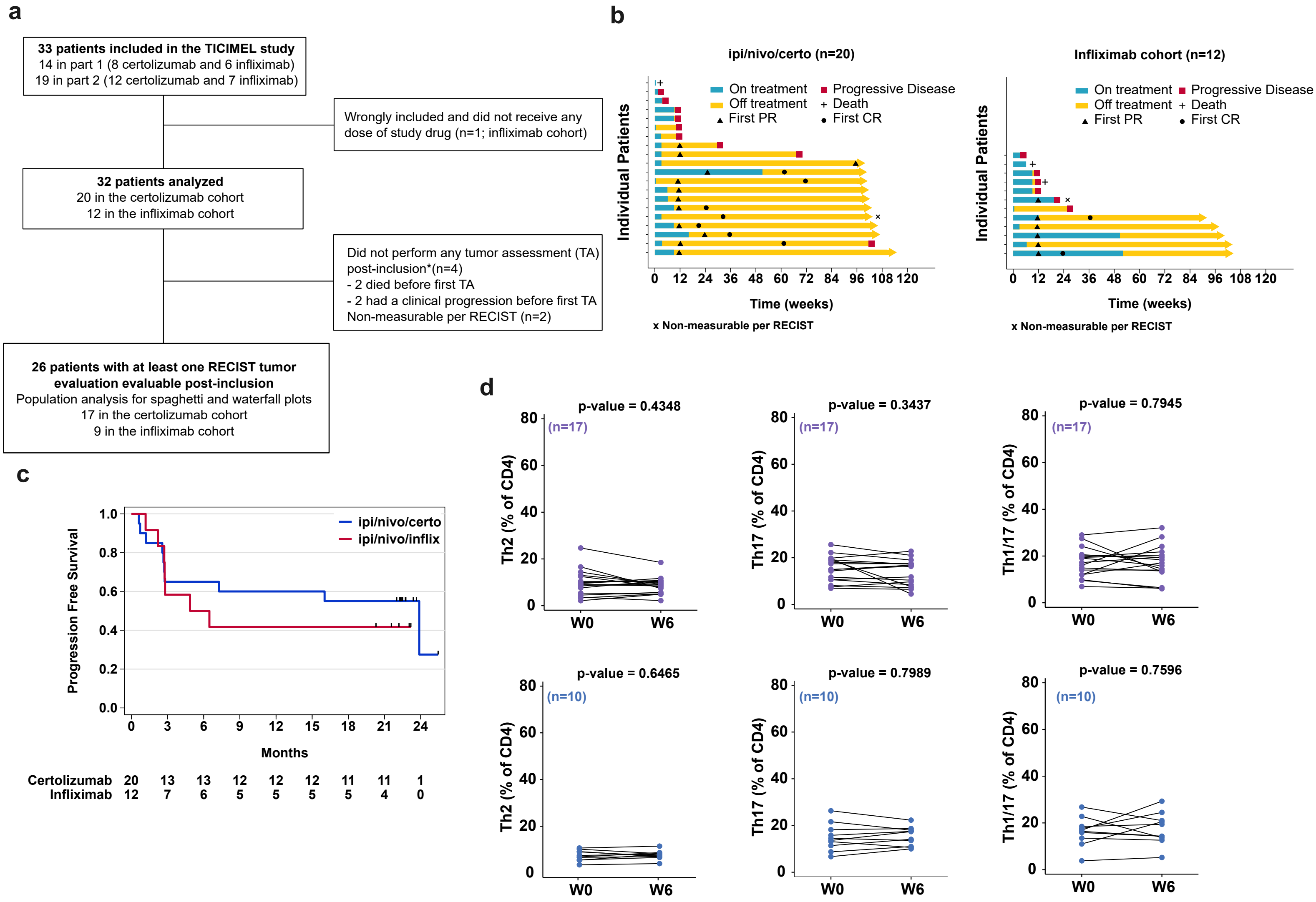


**Extended data 1**

### Supplementary Figure 2

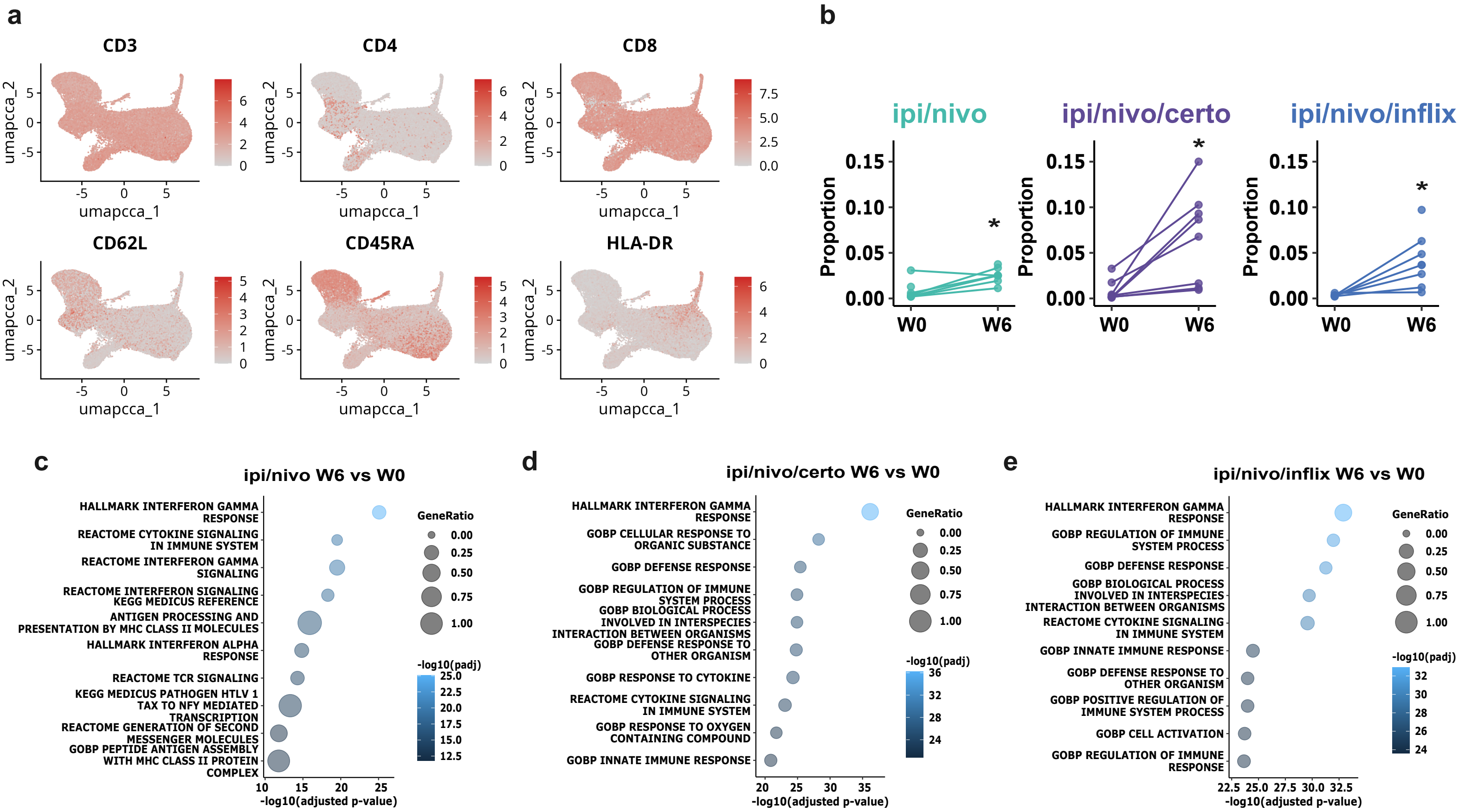


**Extended data 2**

### Supplementary Figure 3

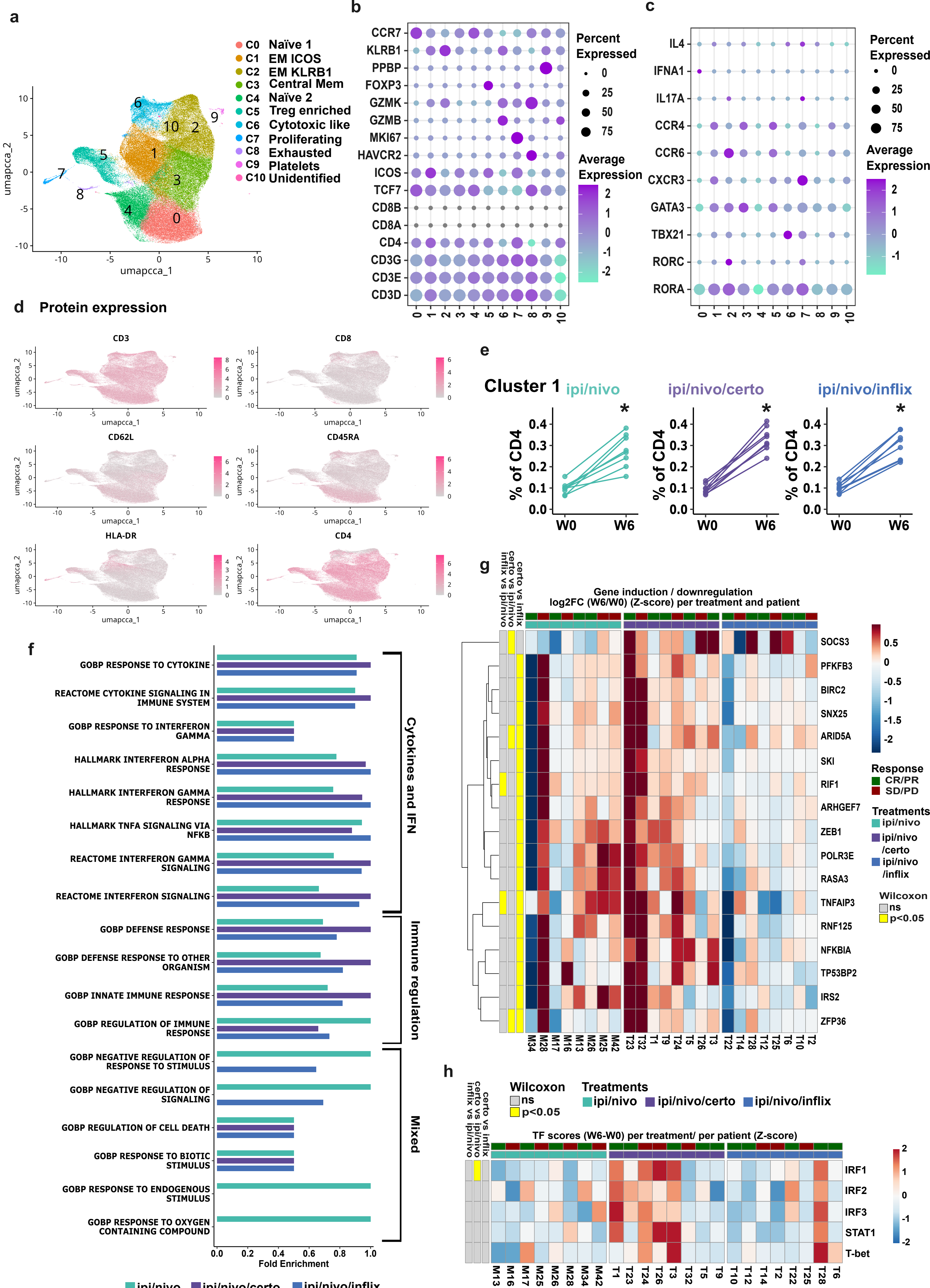


Extended data 3

### Supplementary Figure 4

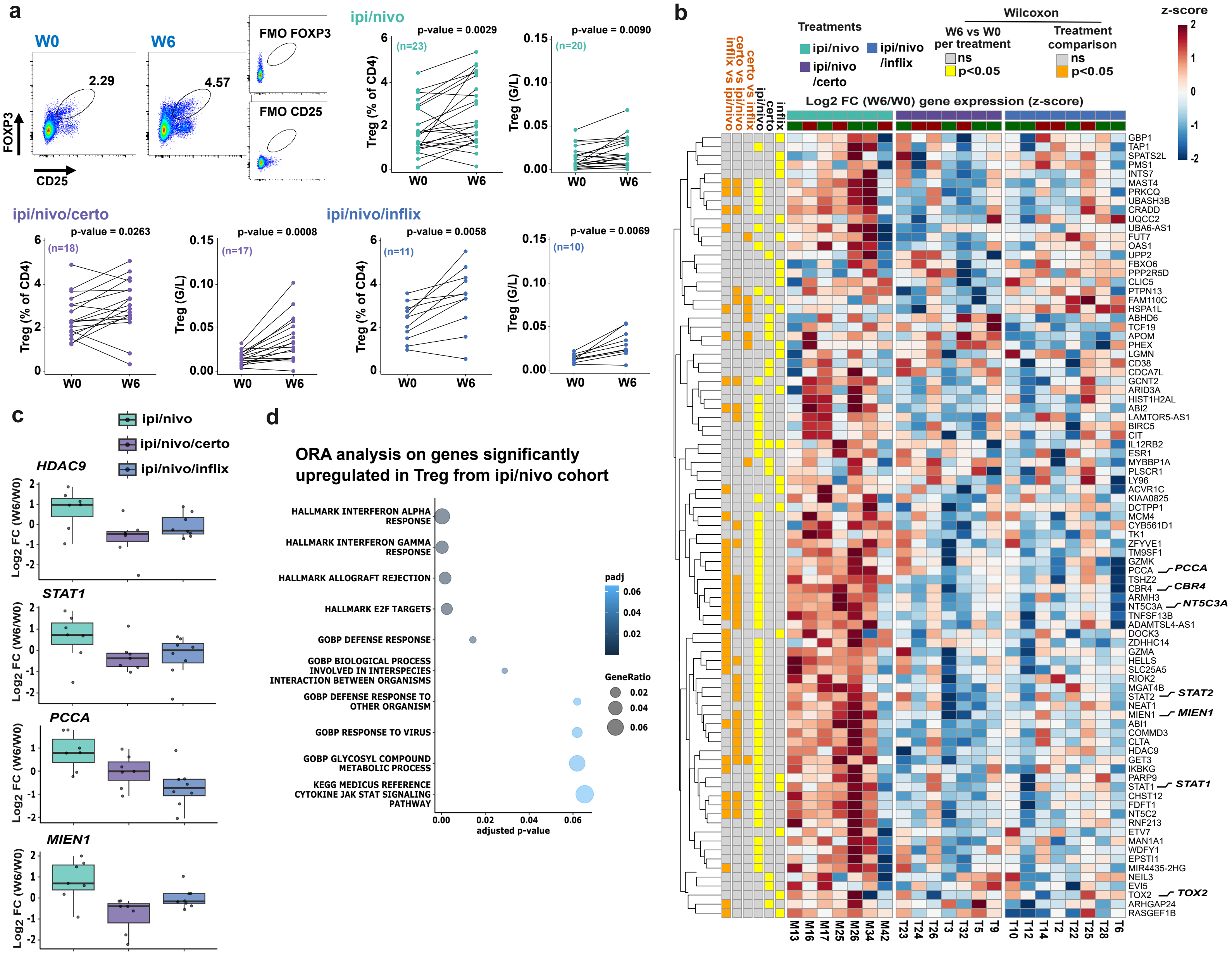


**Extended data 4**

### Supplementary Figure 5

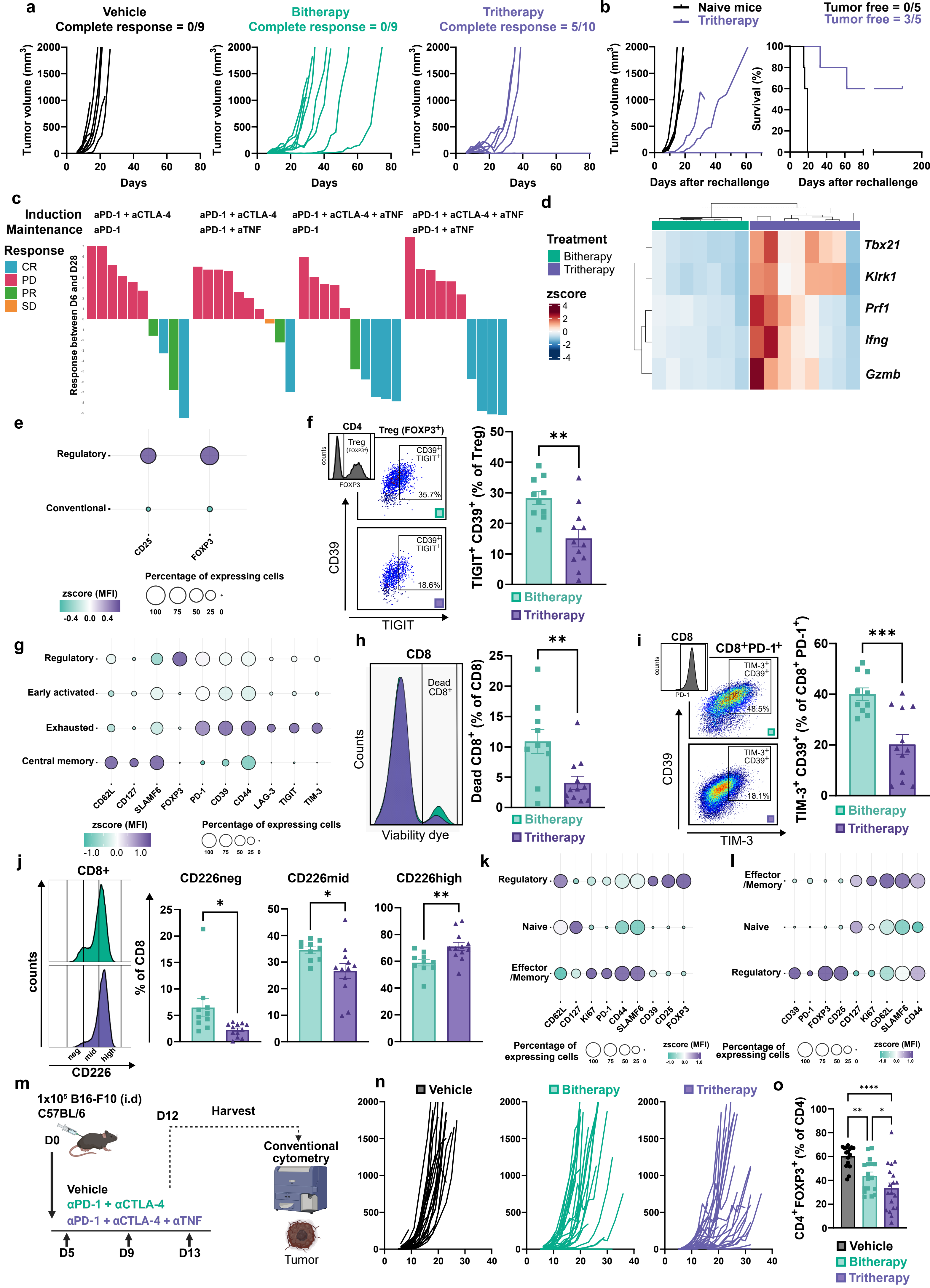


Extended data 5

### Supplementary Figure 6

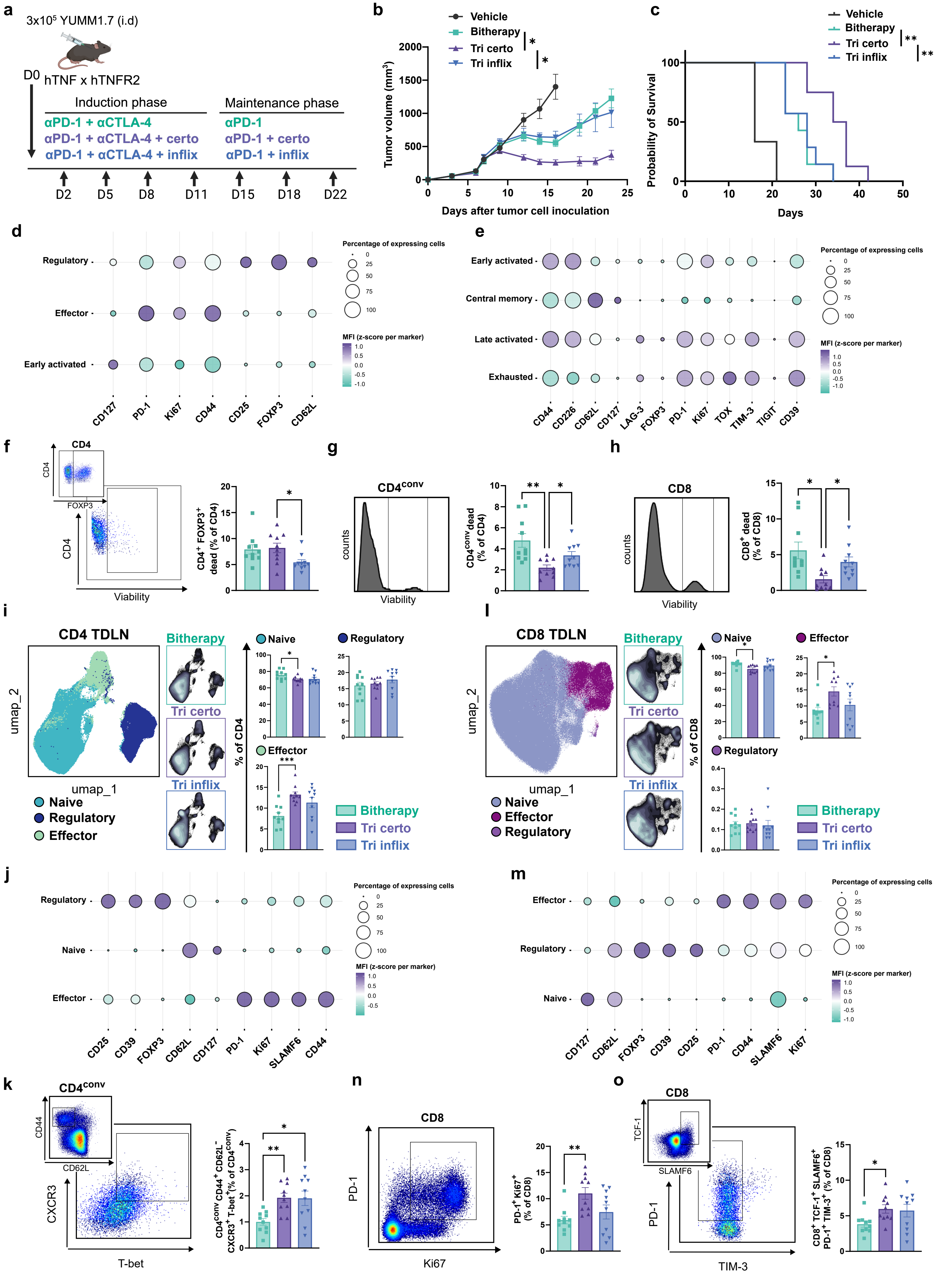


**Extended data 6**

### Supplementary Figure 7

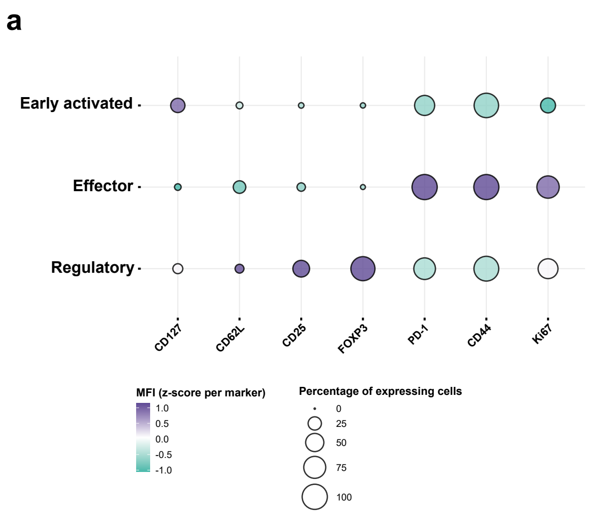


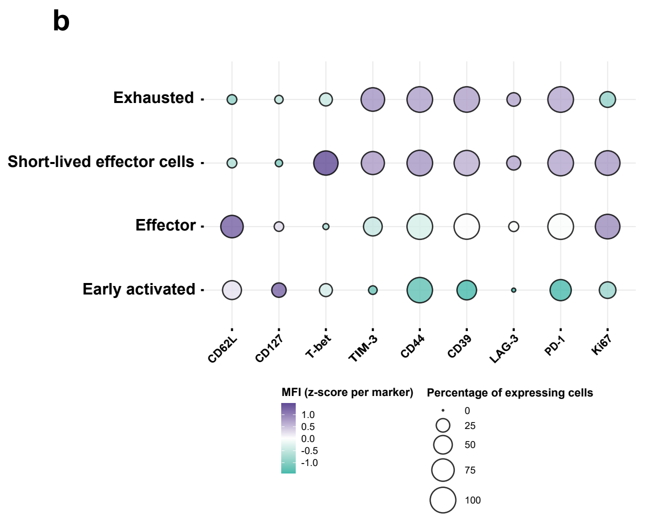


**Extended data 7**
